# Neurometabolite changes in response to antidepressant medication: a systematic review of ^1^H-MRS findings

**DOI:** 10.1101/2023.06.16.23291487

**Authors:** Daphne E. Boucherie, Liesbeth Reneman, Henricus G. Ruhé, Anouk Schrantee

## Abstract

Selective serotonin reuptake inhibitors (SSRIs), serotonin and noradrenaline reuptake inhibitors (SNRIs), and (es-)ketamine are used to treat major depressive disorder (MDD). These different types of medication may involve common neural pathways related to glutamatergic and GABAergic neurotransmitter systems, both of which have been implicated in MDD pathology. We conducted a systematic review of pharmacological ^1^H-MRS studies in healthy volunteers and MDD patients to explore the potential impact of these medications on glutamatergic and GABAergic systems. Studies on SSRIs and SNRIs were highly variable, generally underpowered, and yielded no consistent findings across brain regions or specific populations. Although studies on (es-)ketamine were also highly variable, some demonstrated an increase in glutamate levels in the anterior cingulate cortex in a time-dependent manner after administration. Our findings highlight the need for standardized study and acquisition protocols. Additionally, measuring metabolites dynamically over time or combining ^1^H-MRS with whole brain functional imaging techniques could provide valuable insights into the effects of these medications on glutamate and GABAergic neurometabolism.

## 1. INTRODUCTION

Major depressive disorder (MDD) is a debilitating and highly prevalent psychiatric disorder. Selective serotonin reuptake inhibitors (SSRIs) and serotonin and noradrenaline reuptake inhibitors (SNRIs) are first-choice classes of antidepressants, commonly prescribed for the treatment of MDD and other mood disorders. Although the mechanism of action of SSRIs is not fully understood, they selectively block the reuptake of serotonin from the synaptic cleft, whereas SNRIs additionally inhibit the noradrenaline transporter. This results in increased levels of serotonin (and noradrenaline) in the brain, which is thought to be associated with their antidepressant properties. While SSRIs and SNRIs are generally considered safe and effective, not all patients respond to treatment with these types of medication (Al-Harbi, 2012).

(Es-)Ketamine is another type of antidepressant medication that has gained attention in recent years due to its rapid onset of action and effectiveness in treatment-resistant depression (TRD; Berman *et al*., 2000; Zarate *et al*., 2006). (Es-)Ketamine is a dissociative anesthetic that works by non-selectively antagonizing the N-methyl-D-aspartate (NMDA) receptor, a glutamatergic receptor in the brain (Orser et al., 1997). These receptors are predominantly situated on GABAergic parvalbumin interneurons (Grunze et al., 1996; Ng et al., 2018). The NMDA antagonism results in increased glutamate release, which is thought to enhance synaptic plasticity and neuroplasticity (Buck et al. 2006; Keilhoff et al. 2004) and has been implicated in (es-)ketamine’s antidepressant effects. Although the exact mechanism of action of (es-)ketamine is not fully understood either, its rapid onset of action and effectiveness in treating TRD have made it a promising avenue for the development of new antidepressant medication.

Alterations in glutamatergic and GABAergic systems, through downstream or direct engagement, have been proposed as a potential overlapping mechanism of SSRI, SNRIs and (es-)ketamine antidepressant medications (Skolnick et al., 1996). Indeed, results from animal, post-mortem, imaging, pharmacological, and genetic studies linking alterations in glutamatergic and GABAergic systems to the pathology of MDD have led to the glutamate hypothesis of depression, which suggests an altered glutamatergic metabolism as a mediator of MDD pathology (Sanacora et al. 2012). Interestingly, proton Magnetic Resonance Spectroscopy (^1^H-MRS), a non-invasive neuroimaging technique that enables direct measurements of metabolite levels *in vivo*, has provided evidence of reduced glutamatergic metabolites and lower GABA levels in patients with MDD (Lener et al., 2017; Moriguchi et al., 2019; Yüksel and Öngür, 2010). However, a systematic review of studies about the effects of antidepressant medications on the glutamatergic and GABAergic systems has not been conducted so far.

In this systematic review, we investigate changes in glutamatergic and GABAergic neurotransmission induced by three types of antidepressant medications: SSRIs, SNRIs, and (es-)ketamine. To this purpose, we collected and analyzed human (pharmacological) ^1^H-MRS findings to summarize all available evidence on changes in glutamate, glutamine, glutamate+glutamine (Glx), and GABA in response to acute administration of, or treatment with these medications. Our analysis aims to determine if alterations in these neurotransmitter systems represent a shared biological pathway between different types of antidepressant medication and their response in MDD. Firstly, we evaluate studies that assess the effect of antidepressants on metabolite concentrations in healthy volunteers and MDD patients. Secondly, we investigate the relation between metabolite concentrations and clinical outcomes.

## 2. METHODS

This systematic review was conducted in accordance with the Preferred Reporting Items For Systematic Reviews and Meta-analyses (PRISMA) reporting guideline (Moher et al., 2009). The review protocol was registered in the international prospective register of systematic reviews (PROSPERO; CRD42022384696).

### 2.1 Search strategy

PubMed, Web of Science and Embase were searched from inception to January 10^th^ 2023. See Supplementary Table 1 (Supplementary Methods) for the full search terms and results per database. All titles and abstracts of retrieved publications were independently screened by two researchers (DB and AS) to assess eligibility for inclusion. In case of inconsistencies, full-text articles were obtained. After this initial eligibility check, full text articles were independently screened by AS and DB based on inclusion criteria below. Furthermore, reference lists of selected articles were screened for potential additional studies.

### 2.2 Inclusion criteria

Studies were included when 1) the study design was a randomized controlled trial or a longitudinal cohort study with pre- and post-measurements, 2) the study assessed the effects of SSRIs, SNRIs, or (es-)ketamine on neurometabolism, 3) used ^1^H-MRS or Magnetic Resonance Spectroscopic Imaging (MRSI) to assess metabolite levels, 4) healthy volunteers or subjects primarily diagnosed with MDD were studied (studies including MDD patients with comorbid psychiatric disorders were not excluded). Additionally, studies were included when 5) glutamate, glutamine, Glx or GABA were amongst the investigated metabolites, 6) studies used a field strength >1.5 T, and 7) when the analysis consisted of comparison of metabolite levels between antidepressant treatment and placebo or when comparing baseline with post-medication metabolite levels. We excluded animal studies, duplicate publications, or articles in which the ^1^H-MRS data had already been described in another included article.

### 2.3 Data extraction

Data extraction was done independently by DB and AS using the same summary table with pre-specified fields. We identified demographic and clinical characteristics: the number of subjects, healthy volunteers versus MDD patients, medication status, medication administration (i.e. medication type, route of administration, dose and duration), experimental design, ^1^H-MRS parameters (e.g. field strength, sequence and voxel size), voxel placement, metabolites and quantification method (reference metabolite and software) and symptom severity measures. For the articles investigating the effect of SSRI and SNRI administration or treatment, the duration was classified as acute (single dose), subchronic (≤4 weeks), or chronic (>4 weeks). Results were described for healthy volunteers and MDD patients separately.

### 2.4 Quality assessment of included studies

The NIH quality assessment tools for controlled interventions and for before-after studies with no control group were used (https://www.nhlbi.nih.gov/health-topics/study-quality-assessment-tools). These measures can be used to assess the risk of bias for each of the included studies. We additionally assessed the quality of MRS-reporting using the MRS-Q assessment tool (Peek et al. 2020; and https://osf.io/8s7j9), a new quality appraisal tool based on consensus papers and expert opinion on best-practice. Studies were quantified as poor, fair, or good based on the NIH quality assessment tools.

## 3. RESULTS

The PRISMA flowchart (Figure 1) summarizes study selection (Moher et al., 2009). Twenty-seven studies met the above-mentioned in- and exclusion criteria. Additionally, two articles were included after screening the references lists of included articles, yielding a total of 29 included articles. Extracted features are shown in table 1 and 2 and findings are visualized in Figure 2 and 3.

**Figure 1.**
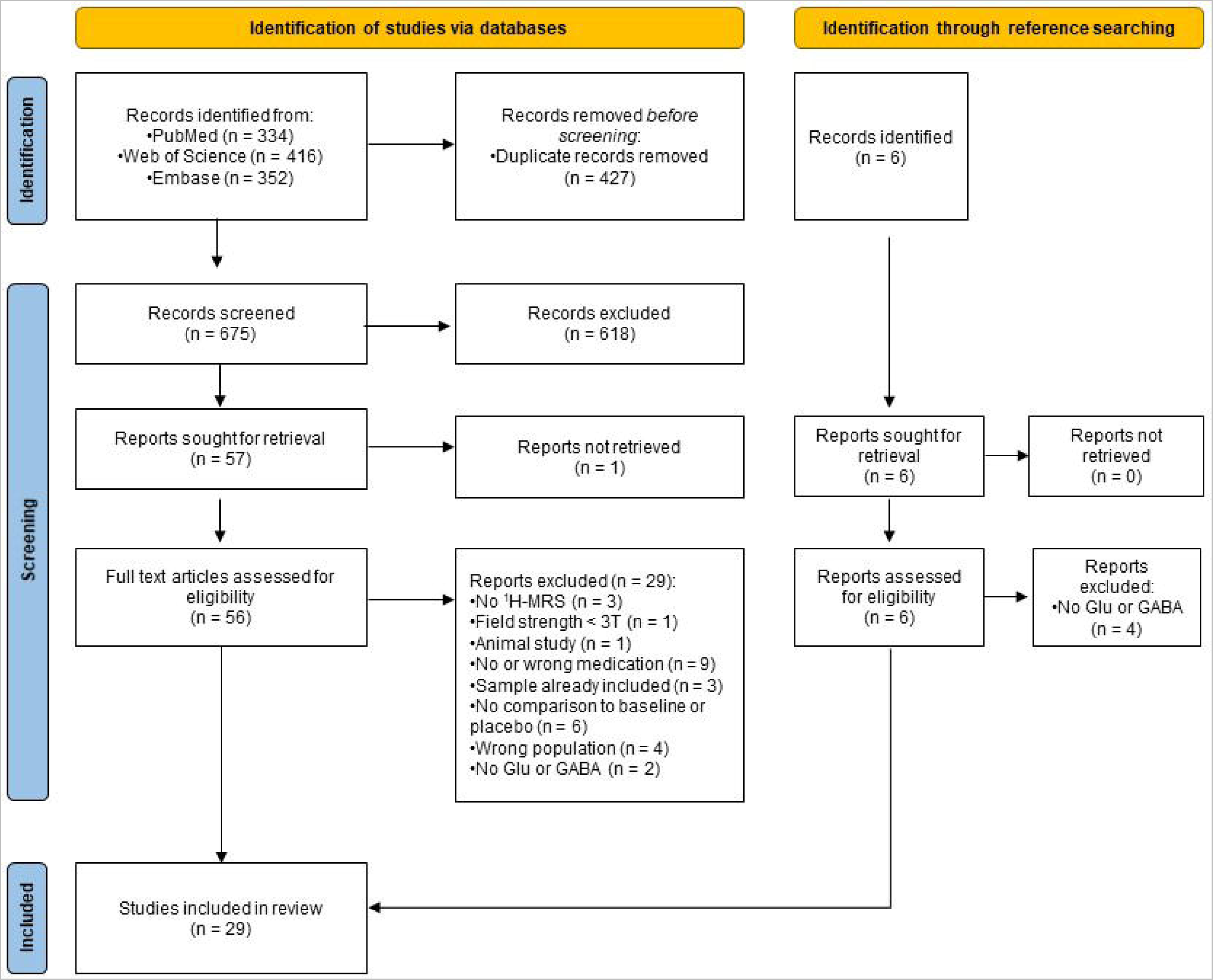
Literature PRISMA flowchart.

**Table 1.**
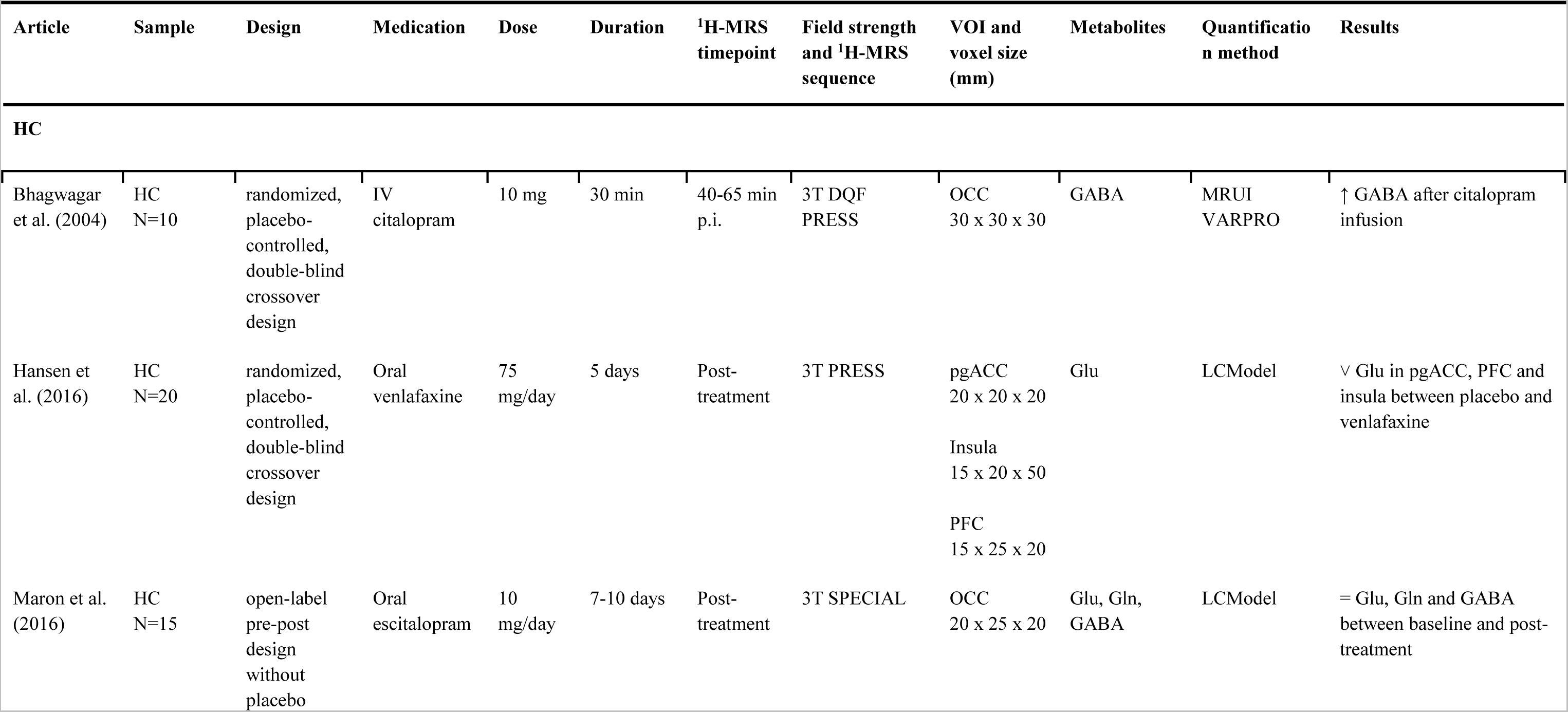
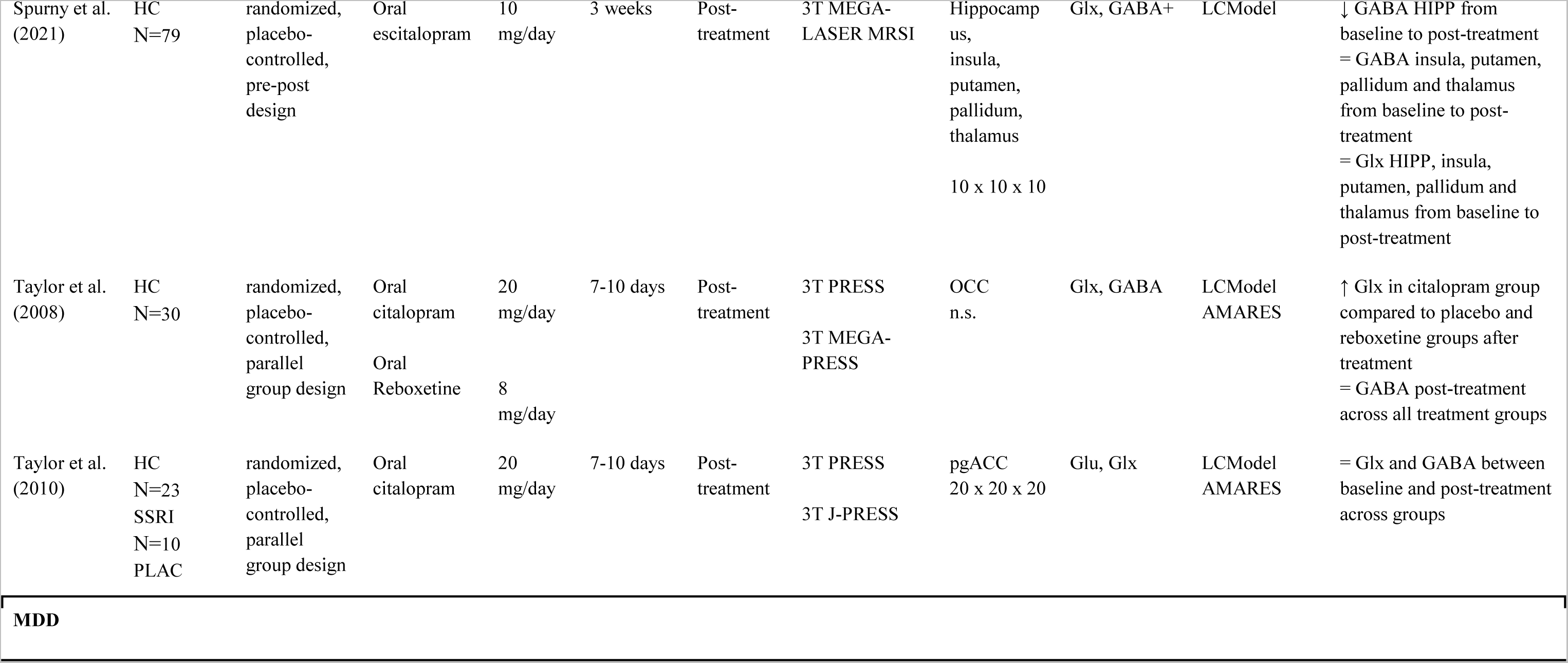
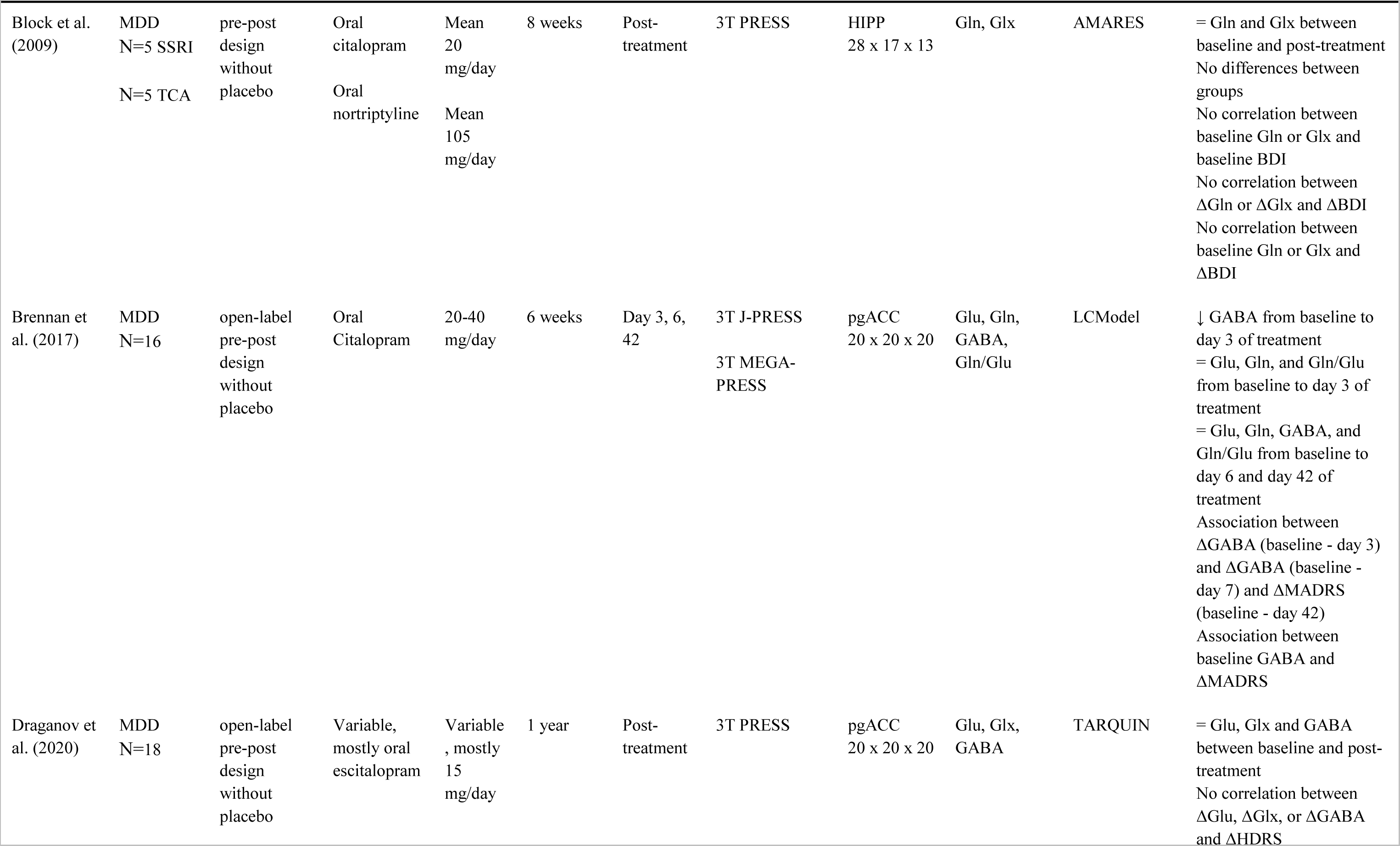
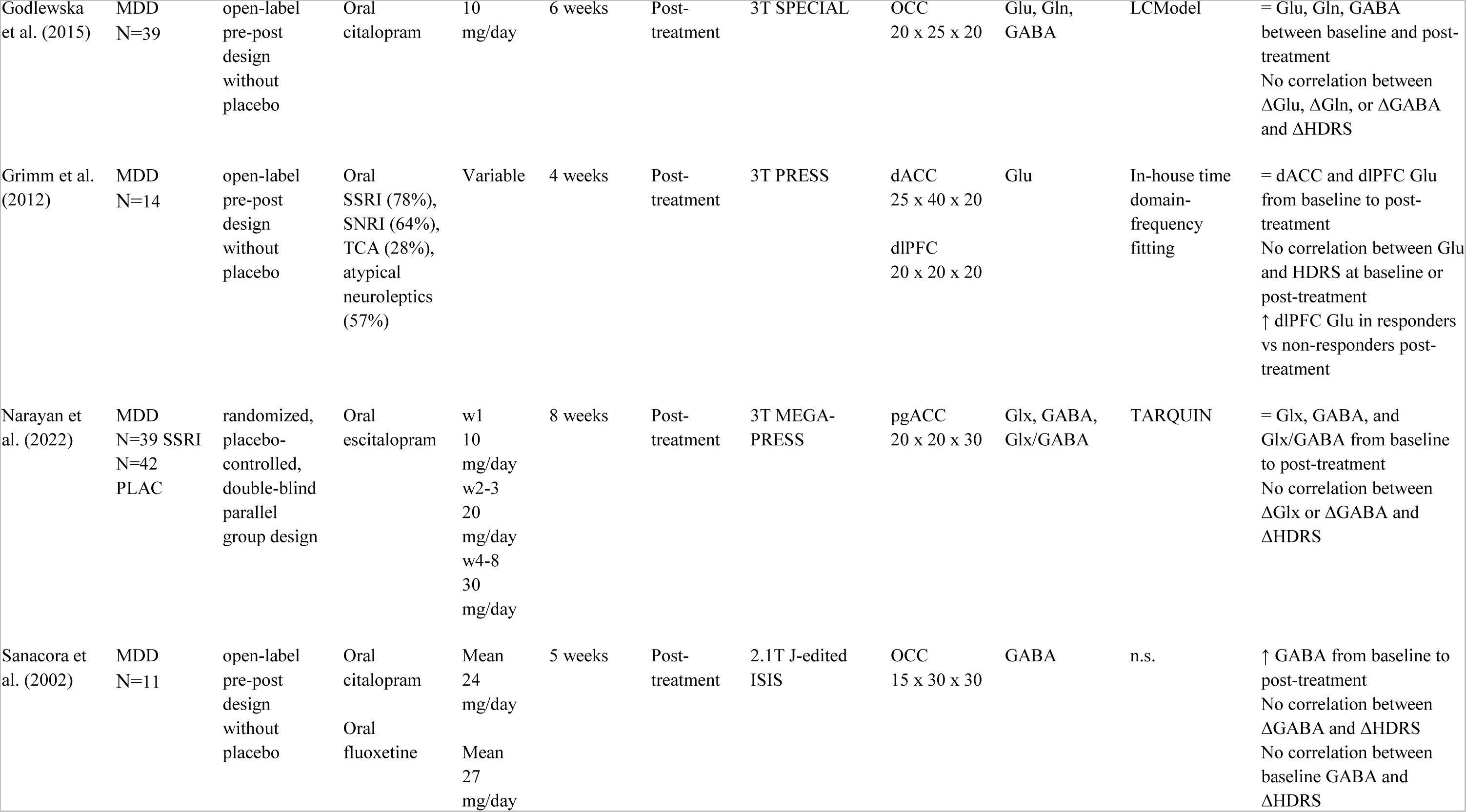
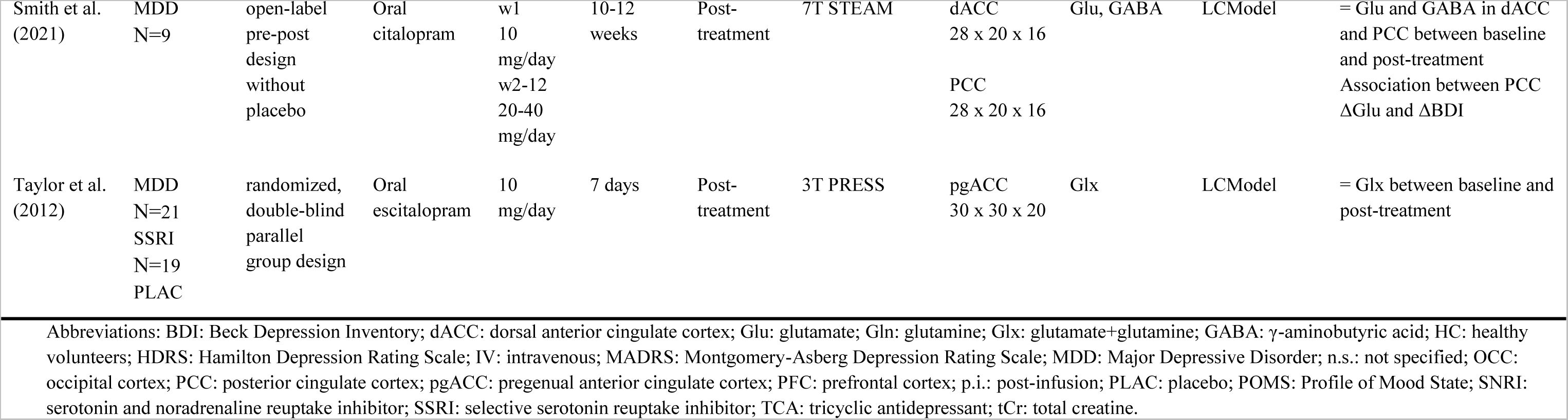
Outcomes of included studies investigating the effect of SSRI or SNRI administration or treatment on glutamatergic or GABAergic metabolites.

| Article | Sample | Design | Medication | Dose | Duration | <sup>1</sup>H-MRS<br>timepoint | Field strength<br>and <sup>1</sup>H-MRS<br>sequence | VOI and<br>voxel size<br>(mm) | Metabolites | Quantification<br>method | Results |
| --- | --- | --- | --- | --- | --- | --- | --- | --- | --- | --- | --- |
| **HC** |  |  |  |  |  |  |  |  |  |  |  |
| Bhagwagar<br>et al. (2004) | HC<br>N=10 | randomized,<br>placebo-controlled,<br>double-blind<br>crossover<br>design | IV<br>citalopram | 10 mg | 30 min | 40-65 min<br>p.i. | 3T DQF<br>PRESS | OCC<br>30 x 30 x 30 | GABA | MRUI<br>VARPRO | ↑ GABA after citalopram<br>infusion |
| Hansen et<br>al. (2016) | HC<br>N=20 | randomized,<br>placebo-controlled,<br>double-blind<br>crossover<br>design | Oral<br>venlafaxine | 75<br>mg/day | 5 days | Post-treatment | 3T PRESS | pgACC<br>20 x 20 x 20<br>Insula<br>15 x 20 x 50<br>PFC<br>15 x 25 x 20 | Glu | LCModel | ∨ Glu in pgACC, PFC and<br>insula between placebo and<br>venlafaxine |
| Maron et al.<br>(2016) | HC<br>N=15 | open-label<br>pre-post<br>design<br>without<br>placebo | Oral<br>escitalopram | 10<br>mg/day | 7-10 days | Post-treatment | 3T SPECIAL | OCC<br>20 x 25 x 20 | Glu, Gln,<br>GABA | LCModel | = Glu, Gln and GABA<br>between baseline and post-treatment |
| Spurny et al.<br>(2021) | HC<br>N=79 | randomized,<br>placebo-<br>controlled,<br>pre-post<br>design | Oral<br>escitalopram | 10<br>mg/day | 3 weeks | Post-<br>treatment | 3T MEGA-<br>LASER MRSI | Hippocamp<br>us,<br>insula,<br>putamen,<br>pallidum,<br>thalamus<br>10 x 10 x 10 | Glx, GABA+ | LCModel | ↓ GABA HIPP from<br>baseline to post-treatment<br>= GABA insula, putamen,<br>pallidum and thalamus<br>from baseline to post-<br>treatment<br>= Glx HIPP, insula,<br>putamen, pallidum and<br>thalamus from baseline to<br>post-treatment |
| Taylor et al.<br>(2008) | HC<br>N=30 | randomized,<br>placebo-controlled,<br>parallel<br>group design | Oral<br>citalopram<br>Oral<br>Reboxetine | 20<br>mg/day<br>8<br>mg/day | 7-10 days | Post-treatment | 3T PRESS<br>3T MEGA-<br>PRESS | OCC<br>n.s. | Glx, GABA | LCModel<br>AMARES | ↑ Glx in citalopram group<br>compared to placebo and<br>reboxetine groups after<br>treatment<br>= GABA post-treatment<br>across all treatment groups |
| Taylor et al.<br>(2010) | HC<br>N=23<br>SSRI<br>N=10<br>PLAC | randomized,<br>placebo-controlled,<br>parallel<br>group design | Oral<br>citalopram | 20<br>mg/day | 7-10 days | Post-treatment | 3T PRESS<br>3T J-PRESS | pgACC<br>20 x 20 x 20 | Glu, Glx | LCModel<br>AMARES | = Glx and GABA between<br>baseline and post-treatment<br>across groups |
| **MDD** |  |  |  |  |  |  |  |  |  |  |  |
| Block et al.<br>(2009) | MDD<br>N=5 SSRI<br>N=5 TCA | pre-post<br>design<br>without<br>placebo | Oral<br>citalopram<br>Oral<br>nortriptyline | Mean<br>20<br>mg/day<br>Mean<br>105<br>mg/day | 8 weeks | Post-treatment | 3T PRESS | HIPP<br>28 x 17 x 13 | Gln, Glx | AMARES | = Gln and Glx between<br>baseline and post-treatment<br>No differences between<br>groups<br>No correlation between<br>baseline Gln or Glx and<br>baseline BDI<br>No correlation between<br>ΔGln or ΔGlx and ΔBDI<br>No correlation between<br>baseline Gln or Glx and<br>ΔBDI |
| Brennan et<br>al. (2017) | MDD<br>N=16 | open-label<br>pre-post<br>design<br>without<br>placebo | Oral<br>Citalopram | 20-40<br>mg/day | 6 weeks | Day 3, 6,<br>42 | 3T J-PRESS<br>3T MEGA-<br>PRESS | pgACC<br>20 x 20 x 20 | Glu, Gln,<br>GABA,<br>Gln/Glu | LCModel | ↓ GABA from baseline to<br>day 3 of treatment<br>= Glu, Gln, and Gln/Glu<br>from baseline to day 3 of<br>treatment<br>= Glu, Gln, GABA, and<br>Gln/Glu from baseline to<br>day 6 and day 42 of<br>treatment<br>Association between<br>ΔGABA (baseline - day 3)<br>and ΔGABA (baseline -<br>day 7) and ΔMADRS<br>(baseline - day 42)<br>Association between<br>baseline GABA and<br>ΔMADRS |
| Draganov et<br>al. (2020) | MDD<br>N=18 | open-label<br>pre-post<br>design<br>without<br>placebo | Variable,<br>mostly oral<br>escitalopram | Variable<br>, mostly<br>15<br>mg/day | 1 year | Post-treatment | 3T PRESS | pgACC<br>20 x 20 x 20 | Glu, Glx,<br>GABA | TARQUIN | = Glu, Glx and GABA<br>between baseline and post-treatment<br>No correlation between<br>ΔGlu, ΔGlx, or ΔGABA<br>and ΔHDRS |
| Godlewska et al. (2015) | MDD<br>N=39 | open-label<br>pre-post<br>design<br>without<br>placebo | Oral<br>citalopram | 10<br>mg/day | 6 weeks | Post-treatment | 3T SPECIAL | OCC<br>20 x 25 x 20 | Glu, Gln,<br>GABA | LCModel | = Glu, Gln, GABA<br>between baseline and post-treatment<br>No correlation between<br>ΔGlu, ΔGln, or ΔGABA<br>and ΔHDRS |
| Grimm et al. (2012) | MDD<br>N=14 | open-label<br>pre-post<br>design<br>without<br>placebo | Oral<br>SSRI (78%),<br>SNRI (64%),<br>TCA (28%),<br>atypical<br>neuroleptics<br>(57%) | Variable | 4 weeks | Post-treatment | 3T PRESS | dACC<br>25 x 40 x 20<br>dlPFC<br>20 x 20 x 20 | Glu | In-house time<br>domain-<br>frequency<br>fitting | = dACC and dlPFC Glu<br>from baseline to post-treatment<br>No correlation between Glu<br>and HDRS at baseline or<br>post-treatment<br>↑ dlPFC Glu in responders<br>vs non-responders post-treatment |
| Narayan et al. (2022) | MDD<br>N=39 SSRI<br>N=42<br>PLAC | randomized,<br>placebo-controlled,<br>double-blind<br>parallel<br>group design | Oral<br>escitalopram | w1<br>10<br>mg/day<br>w2-3<br>20<br>mg/day<br>w4-8<br>30<br>mg/day | 8 weeks | Post-treatment | 3T MEGA-<br>PRESS | pgACC<br>20 x 20 x 30 | Glx, GABA,<br>Glx/GABA | TARQUIN | = Glx, GABA, and<br>Glx/GABA from baseline<br>to post-treatment<br>No correlation between<br>ΔGlx or ΔGABA and<br>ΔHDRS |
| Sanacora et al. (2002) | MDD<br>N=11 | open-label<br>pre-post<br>design<br>without<br>placebo | Oral<br>citalopram<br>Oral<br>fluoxetine | Mean<br>24<br>mg/day<br>Mean<br>27<br>mg/day | 5 weeks | Post-treatment | 2.1T J-edited<br>ISIS | OCC<br>15 x 30 x 30 | GABA | n.s. | ↑ GABA from baseline to<br>post-treatment<br>No correlation between<br>ΔGABA and ΔHDRS<br>No correlation between<br>baseline GABA and<br>ΔHDRS |
| Smith et al.<br>(2021) | MDD<br>N=9 | open-label<br>pre-post<br>design<br>without<br>placebo | Oral<br>citalopram | w1<br>10<br>mg/day<br>w2-12<br>20-40<br>mg/day | 10-12<br>weeks | Post-treatment | 7T STEAM | dACC<br>28 x 20 x 16<br>PCC<br>28 x 20 x 16 | Glu, GABA | LCModel | = Glu and GABA in dACC<br>and PCC between baseline<br>and post-treatment<br>Association between PCC<br>ΔGlu and ΔBDI |
| Taylor et al.<br>(2012) | MDD<br>N=21<br>SSRI<br>N=19<br>PLAC | randomized,<br>double-blind<br>parallel<br>group design | Oral<br>escitalopram | 10<br>mg/day | 7 days | Post-treatment | 3T PRESS | pgACC<br>30 x 30 x 20 | Glx | LCModel | = Glx between baseline and<br>post-treatment |

Abbreviations: BDI: Beck Depression Inventory; dACC: dorsal anterior cingulate cortex; Glu: glutamate; Gln: glutamine; Glx: glutamate+glutamine; GABA: $\gamma$ -aminobutyric acid; HC: healthy volunteers; HDRS: Hamilton Depression Rating Scale; IV: intravenous; MADRS: Montgomery-Asberg Depression Rating Scale; MDD: Major Depressive Disorder; n.s.: not specified; OCC: occipital cortex; PCC: posterior cingulate cortex; pgACC: pregenual anterior cingulate cortex; PFC: prefrontal cortex; p.i.: post-infusion; PLAC: placebo; POMS: Profile of Mood State; SNRI: serotonin and noradrenaline reuptake inhibitor; SSRI: selective serotonin reuptake inhibitor; TCA: tricyclic antidepressant; tCr: total creatine.

### 3.1 SSRIs and SNRIs

Fifteen of the included studies investigated the effect of SSRI or SNRI treatment on metabolite levels (Table 1, Figure 2).

**Figure 2.**
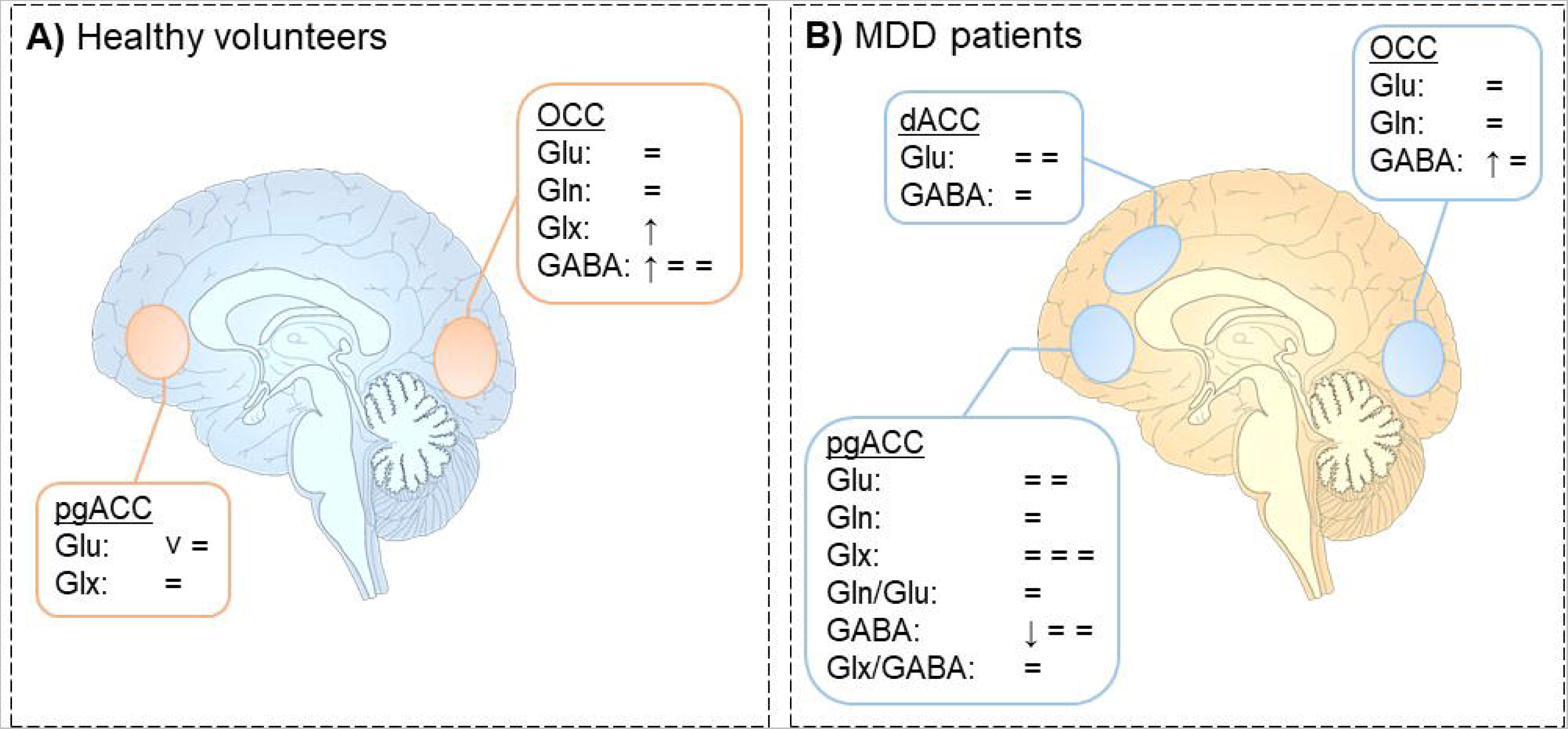
Changes in glutamate, glutamine, Glx and GABA following SSRI or SNRI administration or treatment in a) healthy volunteers and b) MDD patients. Changes are depicted by symbols, with each symbol illustrating one finding. ↑illustrates a significant increase in metabolite concentration; ^ illustrates a trend increase (0.05< p <0.10); ↓ illustrates a significant decrease in metabolite concentration; ៴ illustrates a trend decrease (0.05< p <0.10); = illustrates no changes in metabolite concentrations. dACC: dorsal anterior cingulate cortex; pgACC: pregenual anterior cingulate cortex; OCC: occipital cortex.

#### 3.1.1 SSRIs and SNRIs in healthy volunteers

Six studies investigated the effect of SSRIs and SNRIs on glutamatergic or GABAergic metabolite levels in healthy volunteers, with most studies focusing on either pregenual anterior cingulate cortex (pgACC) (two studies) or the occipital cortex (OCC) (three studies), which are therefore discussed separately (Table 1, Figure 2a). All studies administered medication orally, except for one (Bhagwagar et al., 2004).

##### 3.1.1.1 Pregenual anterior cingulate cortex

In the first study, no effect of treatment on either Glu or Glx in pgACC was observed in subjects receiving either subchronic citalopram (SSRI; 20 mg/day; n=23) or placebo (n=10) for a duration of 7-10 days (Taylor et al., 2010). In contrast, a subchronic 5-day, randomized, placebo-controlled crossover study with venlafaxine (SNRI; 75 mg/day) observed a trend decrease in Glu in the pgACC in 20 subjects (Hansen et al., 2016).

##### 3.1.1.2 Occipital cortex

In a study by Bhagwagar et al. (2004), an acute intravenous citalopram (SSRI; 10 mg; n=10) dose resulted in an increase in occipital GABA between 40-65 minutes after completion of the citalopram infusion. In contrast, Taylor et al. (2008) found no treatment group differences in occipital GABA in 30 participants following 7-10 days of subchronic citalopram (SSRI; 20 mg/day) treatment. However, they observed an increase in Glx for the citalopram group compared to the placebo group. Another study administering escitalopram (SSRI; 10 mg/day) in a similar subchronic design, found no effect of treatment on Glu, Gln and GABA concentrations from baseline to post-treatment in 15 participants (Maron et al., 2016).

##### 3.1.1.3 Other brain regions

Hansen et al. (2016) found a trend decrease in Glu in the PFC and insula after venlafaxine (SNRI; 75 mg/day) treatment. Finally, Spurny *et al*. (2021)investigated Glx and GABA in 79 participants after three weeks of treatment with escitalopram (SSRI; 10 mg/day) using MRSI in five regions (insula, putamen, pallidum, thalamus and hippocampus), during an associative relearning paradigm. They observed a reduction in hippocampal Glx from pre- to post-treatment with escitalopram (SSRI; 10 mg/day), but no effect on GABA. In the putamen, pallidum, and thalamus no differences between baseline and post-treatment were observed in Glx or GABA.

#### 3.1.2 SSRIs and SNRIs in MDD patients

Nine studies assessed the effect of SSRI or SNRI treatment in MDD patients. All patient samples consisted of medication-free patients, except for one study that did not exclude participants based on medication status (Narayan et al., 2022). Some studies used a minimal washout period of 2 or 3 weeks (Brennan et al. 2017; Godlewska et al. 2015; Sanacora et al. 2002; Taylor et al. 2012), whereas another study used a longer washout period of 8 weeks (Block et al., 2009). One study only included patients who had not received medication for one year (Smith et al., 2021) and one study included only patients who were treatment-naive or had minimal exposure (<2 weeks lifelong) to antidepressant treatment (Draganov et al., 2020). The pgACC was assessed four times, the dorsal anterior cingulate cortex (dACC) twice, and the occipital cortex (OCC) also twice (Table 1, Figure 2b).

##### 3.1.2.1 Pregenual anterior cingulate cortex

Taylor et al. (2012) found no differences between subchronic 7-day treatment with escitalopram (SSRI; 10 mg/day; n=21) vs. placebo (n=19) in Glx concentrations in the pgACC. Brennan et al. (2017) investigated the effect of 6 weeks (chronic) open-label citalopram (SSRI; 20-40 mg/day) treatment in 16 MDD patients on Glu, Gln, Gln/Glu and GABA concentrations in the pgACC at multiple time points after treatment onset (at day 3, day 7 and day 42). No differences compared to baseline were observed for Glu, Gln or Gln/Glu for any of these time points. However, they found a decrease in GABA from baseline to day 3 of citalopram treatment. One of the few randomized, placebo-controlled studies (n=39 SSRI and n=41 placebo) observed no differences in Glx or GABA following chronic 8-week treatment with escitalopram (SSRI; 10-30 mg/day) (Narayan et al., 2022). As an additional marker for excitatory-inhibitory balance, the ratio between the two (Glx/GABA) was also investigated, but no changes in this ratio were observed. In a one-year follow-up, Draganov et al. (2020) compared Glu, Glx and GABA levels before and after one year of chronic treatment (variable, mostly SSRI; mostly 10 mg/day) in 18 MDD patients, but also found no significant change over time for any of these metabolites.

##### 3.1.2.2 Dorsal anterior cingulate cortex

An observational study, in which antidepressant medication was not standardized across subjects, assessed Glu levels in the dACC of 14 MDD patients before and after subchronic 4 weeks of treatment (variable; SSRI, SNRI, TCA, atypical neuroleptics, or a combination) (Grimm et al., 2012). They did not observe a time-dependent change in Glu levels. Smith et al. (2021) assessed the effects of chronic 10-12 weeks of citalopram (SSRI; 10-40 mg/day) treatment on dACC Glu and GABA in 9 patients with late-life depression. They likewise did not observe any differences between baseline and post-treatment measurements.

##### 3.1.2.3 Occipital cortex

One study (Sanacora et al., 2002) found increased occipital GABA levels after 5 weeks of chronic citalopram (SSRI; mean 24 mg/day) or fluoxetine (SSRI; mean 27 mg/day) treatment in 11 MDD patients. In contrast, Godlewska et al. (2015) did not find altered concentrations of Glu, Gln or GABA in 39 MDD patients after chronic 6-week citalopram treatment (SSRI; 10 mg/day).

##### 3.1.2.4 Other brain regions

In the PCC, Block et al. (2009) found no group differences and treatment effects of 8-weeks chronic citalopram (SSRI; mean 20 mg/day; n=5) or nortriptyline (TCA; n=5) on hippocampal Glx and Gln. Another study in 9 patients with late-life depression also did not observe an effect of chronic 10-12 weeks citalopram (SSRI; 10-40 mg/day) treatment on PCC Glu or GABA (Smith et al., 2021). In the dlPFC, no changes in response to subchronic 4-week treatment were observed for Glu in 14 MDD patients (Grimm et al., 2012).

#### 3.1.3 Relationship between metabolite measures and symptoms

In the pgACC, Brennan et al. (2017) observed a significant positive association between clinical improvement from baseline to day 42 and change in GABA levels from baseline to day 3 and from baseline to day 7 in 16 MDD patients. Additionally, they observed that lower baseline GABA levels were associated with greater clinical improvement. In contrast, another study found no correlation between Glx, GABA, or Glx/GABA and clinical symptoms, adjusted for treatment type (placebo vs. SSRI), in 81 MDD patients (Narayan et al., 2022). Similarly, the only study conducting a one-year follow-up did not find significant relationships between changes in symptom severity and metabolite levels of Glx, Glu, or GABA over time in 18 MDD patients (Draganov et al., 2020). In the hippocampus, Block et al. (2009) observed no correlation between baseline Glx or Gln with baseline symptom severity score in 10 MDD patients. In the occipital cortex, Sanacora et al. (2002) reported that there was no correlation between the change in occipital GABA and the change in symptom severity score, nor was there a correlation between pre-treatment GABA levels and the change in symptom severity in their sample of 11 MDD patients. Similarly, Godlewska et al. (2015) found no correlation between change in the symptom severity and change in occipital Glu, Gln, or GABA level in 39 MDD patients. Likewise, Grimm et al. (2012) did not find an association in their sample of 14 MDD patients between Glu in the dACC and dlPFC and symptom severity at baseline or after treatment. Finally, (Smith et al., 2021) found that decreases in PCC Glu were associated with improvement in depressive symptoms in their modest sample of 9 MDD patients.

### 3.2 (Es-)Ketamine

Fourteen studies investigated the effect of (es-)ketamine administration on glutamatergic and GABAergic metabolite concentrations (Table 2 and Figure 3).

**Table 2.**
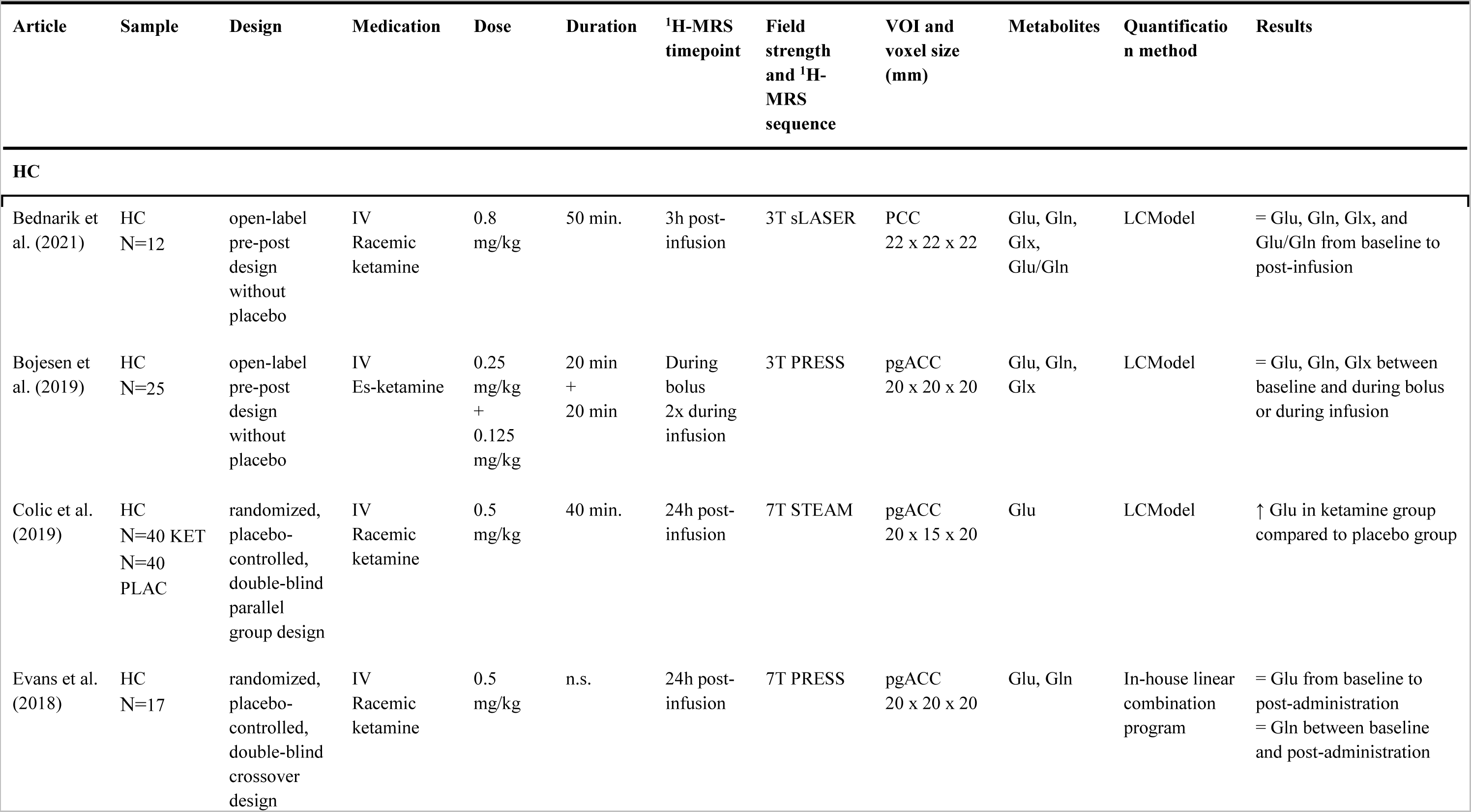
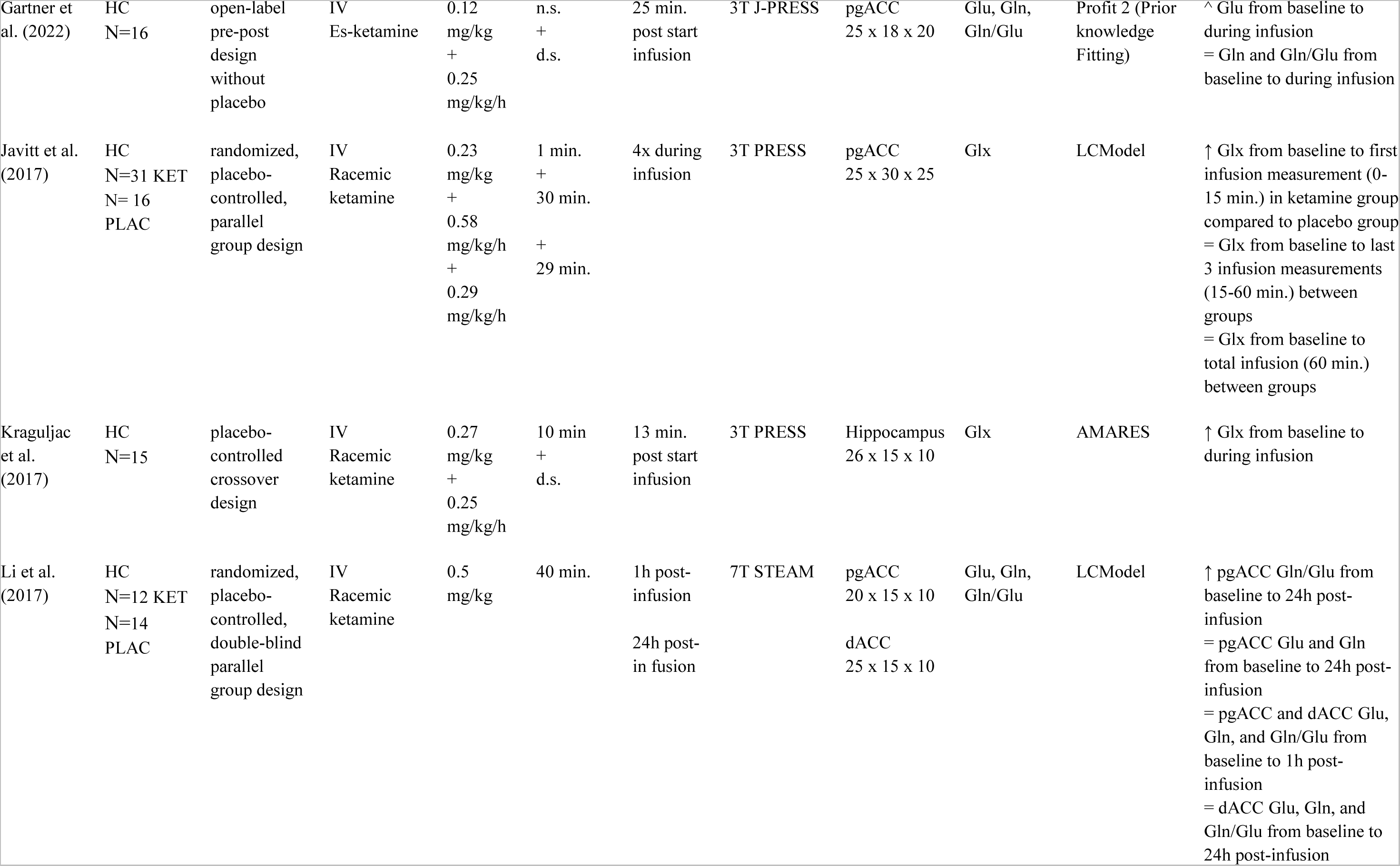
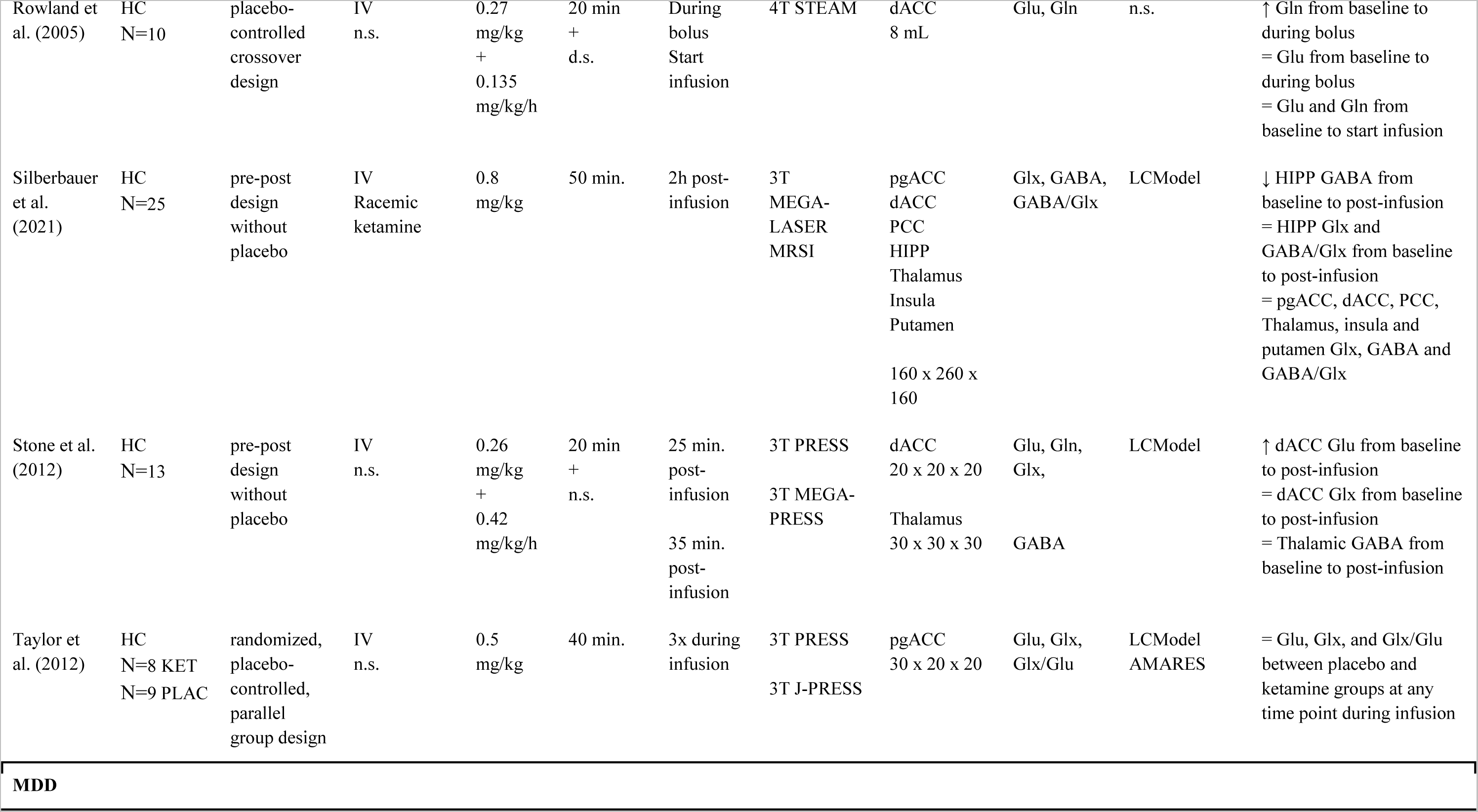
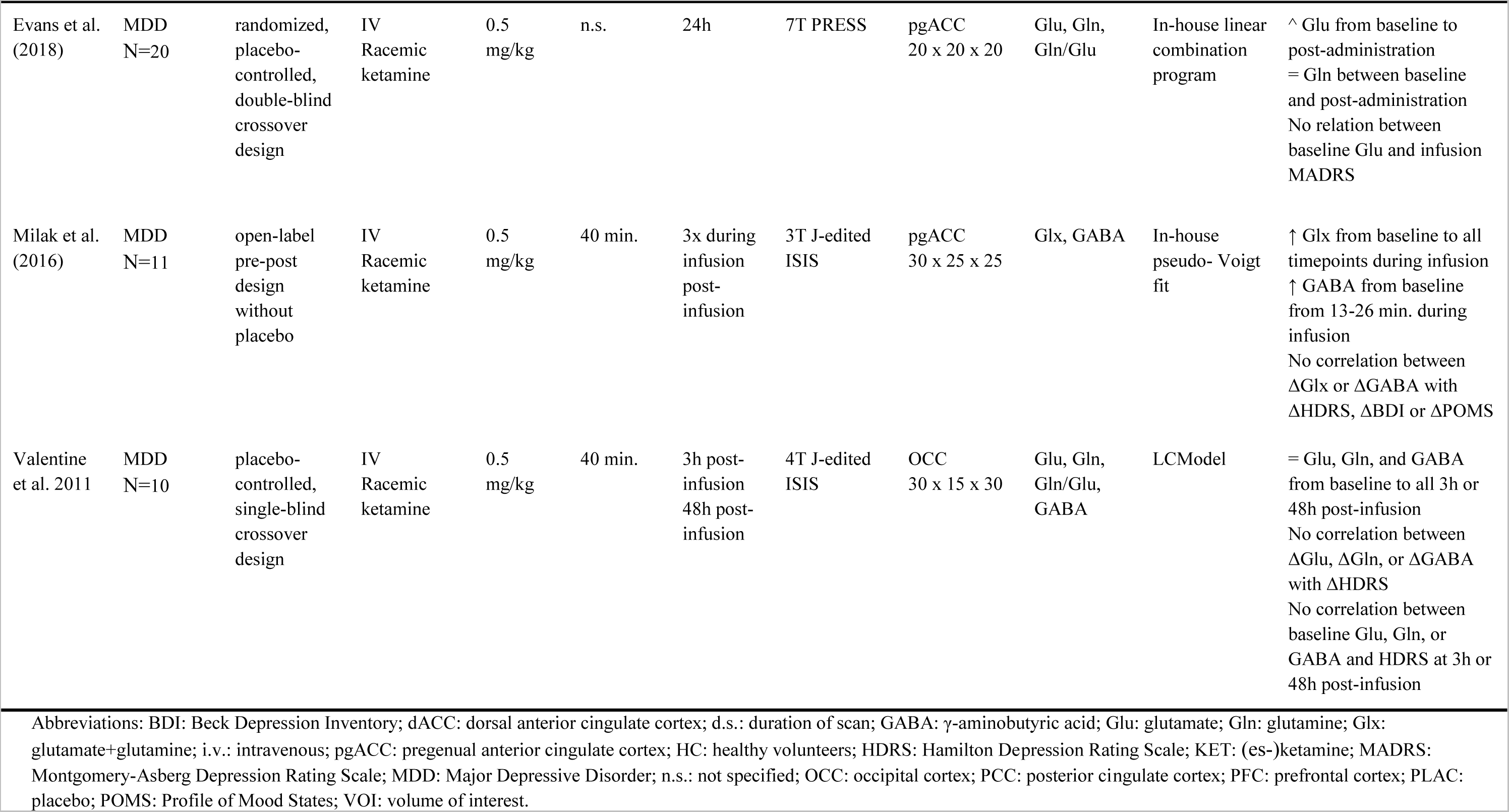
Outcomes of included studies investigating the effect of (es-)ketamine administration on glutamatergic or GABAergic metabolites.

| Article | Sample | Design | Medication | Dose | Duration | <sup>1</sup>H-MRS<br>timepoint | Field<br>strength<br>and <sup>1</sup>H-MRS<br>sequence | VOI and<br>voxel size<br>(mm) | Metabolites | Quantification<br>method | Results |
| --- | --- | --- | --- | --- | --- | --- | --- | --- | --- | --- | --- |
| **HC** |  |  |  |  |  |  |  |  |  |  |  |
| Bednarik et al. (2021) | HC<br>N=12 | open-label<br>pre-post<br>design<br>without<br>placebo | IV<br>Racemic<br>ketamine | 0.8<br>mg/kg | 50 min. | 3h post-infusion | 3T sLASER | PCC<br>22 x 22 x 22 | Glu, Gln,<br>Glx,<br>Glu/Gln | LCModel | = Glu, Gln, Glx, and<br>Glu/Gln from baseline to<br>post-infusion |
| Bojesen et al. (2019) | HC<br>N=25 | open-label<br>pre-post<br>design<br>without<br>placebo | IV<br>Es-ketamine | 0.25<br>mg/kg<br>+<br>0.125<br>mg/kg | 20 min<br>+<br>20 min | During<br>bolus<br>2x during<br>infusion | 3T PRESS | pgACC<br>20 x 20 x 20 | Glu, Gln,<br>Glx | LCModel | = Glu, Gln, Glx between<br>baseline and during bolus<br>or during infusion |
| Colic et al. (2019) | HC<br>N=40 KET<br>N=40<br>PLAC | randomized,<br>placebo-controlled,<br>double-blind<br>parallel<br>group design | IV<br>Racemic<br>ketamine | 0.5<br>mg/kg | 40 min. | 24h post-infusion | 7T STEAM | pgACC<br>20 x 15 x 20 | Glu | LCModel | ↑ Glu in ketamine group<br>compared to placebo group |
| Evans et al. (2018) | HC<br>N=17 | randomized,<br>placebo-controlled,<br>double-blind<br>crossover<br>design | IV<br>Racemic<br>ketamine | 0.5<br>mg/kg | n.s. | 24h post-infusion | 7T PRESS | pgACC<br>20 x 20 x 20 | Glu, Gln | In-house linear<br>combination<br>program | = Glu from baseline to<br>post-administration<br>= Gln between baseline<br>and post-administration |
| Gartner et al. (2022) | HC<br>N=16 | open-label<br>pre-post<br>design<br>without<br>placebo | IV<br>Es-ketamine | 0.12<br>mg/kg<br>+<br>0.25<br>mg/kg/h | n.s.<br>+<br>d.s. | 25 min.<br>post start<br>infusion | 3T J-PRESS | pgACC<br>25 x 18 x 20 | Glu, Gln,<br>Gln/Glu | Profit 2 (Prior<br>knowledge<br>Fitting) | ^ Glu from baseline to<br>during infusion<br>= Gln and Gln/Glu from<br>baseline to during infusion |
| Javitt et al. (2017) | HC<br>N=31 KET<br>N= 16<br>PLAC | randomized,<br>placebo-controlled,<br>parallel<br>group design | IV<br>Racemic<br>ketamine | 0.23<br>mg/kg<br>+<br>0.58<br>mg/kg/h<br>+<br>0.29<br>mg/kg/h | 1 min.<br>+<br>30 min.<br>+<br>29 min. | 4x during<br>infusion | 3T PRESS | pgACC<br>25 x 30 x 25 | Glx | LCModel | ↑ Glx from baseline to first<br>infusion measurement (0-15 min.) in ketamine group<br>compared to placebo group<br>= Glx from baseline to last<br>3 infusion measurements<br>(15-60 min.) between<br>groups<br>= Glx from baseline to<br>total infusion (60 min.)<br>between groups |
| Kraguljac et al. (2017) | HC<br>N=15 | placebo-controlled<br>crossover<br>design | IV<br>Racemic<br>ketamine | 0.27<br>mg/kg<br>+<br>0.25<br>mg/kg/h | 10 min<br>+<br>d.s. | 13 min.<br>post start<br>infusion | 3T PRESS | Hippocampus<br>26 x 15 x 10 | Glx | AMARES | ↑ Glx from baseline to<br>during infusion |
| Li et al. (2017) | HC<br>N=12 KET<br>N=14<br>PLAC | randomized,<br>placebo-controlled,<br>double-blind<br>parallel<br>group design | IV<br>Racemic<br>ketamine | 0.5<br>mg/kg | 40 min. | 1h post-infusion<br>24h post-<br>in fusion | 7T STEAM | pgACC<br>20 x 15 x 10<br>dACC<br>25 x 15 x 10 | Glu, Gln,<br>Gln/Glu | LCModel | ↑ pgACC Gln/Glu from<br>baseline to 24h post-infusion<br>= pgACC Glu and Gln<br>from baseline to 24h post-infusion<br>= pgACC and dACC Glu,<br>Gln, and Gln/Glu from<br>baseline to 1h post-infusion<br>= dACC Glu, Gln, and<br>Gln/Glu from baseline to<br>24h post-infusion |
| Rowland et al. (2005) | HC<br>N=10 | placebo-controlled crossover design | IV<br>n.s. | 0.27 mg/kg + 0.135 mg/kg/h | 20 min + d.s. | During bolus Start infusion | 4T STEAM | dACC<br>8 mL | Glu, Gln | n.s. | ↑ Gln from baseline to during bolus<br>= Glu from baseline to during bolus<br>= Glu and Gln from baseline to start infusion |
| Silberbauer et al. (2021) | HC<br>N=25 | pre-post design without placebo | IV<br>Racemic ketamine | 0.8 mg/kg | 50 min. | 2h post-infusion | 3T MEGA-LASER MRSI | pgACC<br>dACC<br>PCC<br>HIPP<br>Thalamus<br>Insula<br>Putamen<br>160 x 260 x 160 | Glx, GABA, GABA/Glx | LCModel | ↓ HIPP GABA from baseline to post-infusion<br>= HIPP Glx and GABA/Glx from baseline to post-infusion<br>= pgACC, dACC, PCC, Thalamus, insula and putamen Glx, GABA and GABA/Glx |
| Stone et al. (2012) | HC<br>N=13 | pre-post design without placebo | IV<br>n.s. | 0.26 mg/kg + 0.42 mg/kg/h | 20 min + n.s. | 25 min. post-infusion<br>35 min. post-infusion | 3T PRESS<br>3T MEGA-PRESS | dACC<br>20 x 20 x 20<br>Thalamus<br>30 x 30 x 30 | Glu, Gln, Glx,<br>GABA | LCModel | ↑ dACC Glu from baseline to post-infusion<br>= dACC Glx from baseline to post-infusion<br>= Thalamic GABA from baseline to post-infusion |
| Taylor et al. (2012) | HC<br>N=8 KET<br>N=9 PLAC | randomized, placebo-controlled, parallel group design | IV<br>n.s. | 0.5 mg/kg | 40 min. | 3x during infusion | 3T PRESS<br>3T J-PRESS | pgACC<br>30 x 20 x 20 | Glu, Glx, Glx/Glu | LCModel<br>AMARES | = Glu, Glx, and Glx/Glu between placebo and ketamine groups at any time point during infusion |
| MDD |  |  |  |  |  |  |  |  |  |  |  |
| Evans et al. (2018) | MDD<br>N=20 | randomized, placebo-controlled, double-blind crossover design | IV<br>Racemic ketamine | 0.5 mg/kg | n.s. | 24h | 7T PRESS | pgACC<br>20 x 20 x 20 | Glu, Gln, Gln/Glu | In-house linear combination program | ^ Glu from baseline to post-administration = Gln between baseline and post-administration<br>No relation between baseline Glu and infusion MADRS |
| Milak et al. (2016) | MDD<br>N=11 | open-label pre-post design without placebo | IV<br>Racemic ketamine | 0.5 mg/kg | 40 min. | 3x during infusion post-infusion | 3T J-edited ISIS | pgACC<br>30 x 25 x 25 | Glx, GABA | In-house pseudo- Voigt fit | ↑ Glx from baseline to all timepoints during infusion<br>↑ GABA from baseline from 13-26 min. during infusion<br>No correlation between ΔGlx or ΔGABA with ΔHDRS, ΔBDI or ΔPOMS |
| Valentine et al. 2011 | MDD<br>N=10 | placebo-controlled, single-blind crossover design | IV<br>Racemic ketamine | 0.5 mg/kg | 40 min. | 3h post-infusion<br>48h post-infusion | 4T J-edited ISIS | OCC<br>30 x 15 x 30 | Glu, Gln, Gln/Glu, GABA | LCModel | = Glu, Gln, and GABA from baseline to all 3h or 48h post-infusion<br>No correlation between ΔGlu, ΔGln, or ΔGABA with ΔHDRS<br>No correlation between baseline Glu, Gln, or GABA and HDRS at 3h or 48h post-infusion |

Abbreviations: BDI: Beck Depression Inventory; dACC: dorsal anterior cingulate cortex; d.s.: duration of scan; GABA: $\gamma$ -aminobutyric acid; Glu: glutamate; Gln: glutamine; Glx: glutamate+glutamine; i.v.: intravenous; pgACC: pregenual anterior cingulate cortex; HC: healthy volunteers; HDRS: Hamilton Depression Rating Scale; KET: (es-)ketamine; MADRS: Montgomery-Asberg Depression Rating Scale; MDD: Major Depressive Disorder; n.s.: not specified; OCC: occipital cortex; PCC: posterior cingulate cortex; PFC: prefrontal cortex; PLAC: placebo; POMS: Profile of Mood States; VOI: volume of interest.

**Figure 3.**
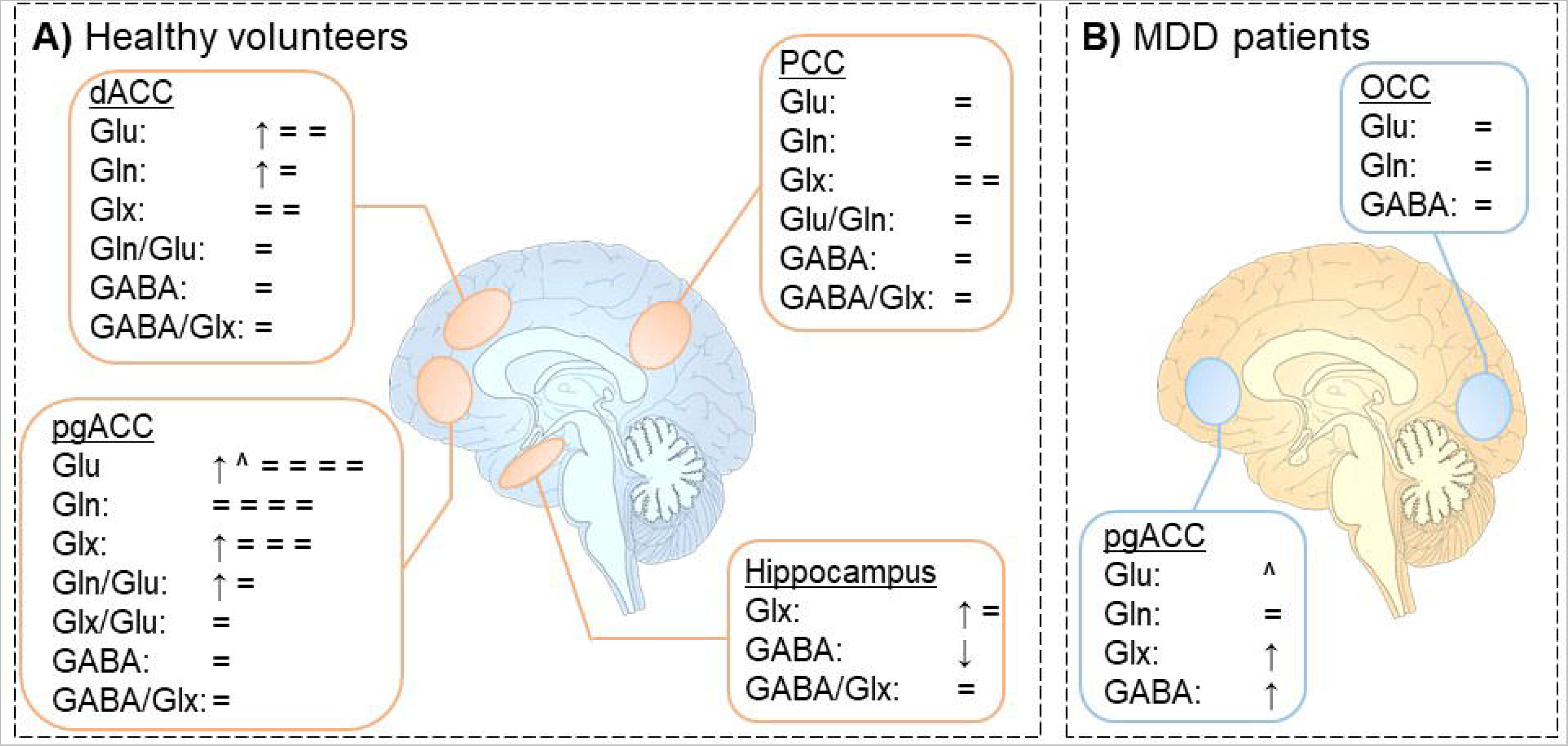
Changes in glutamate, glutamine, Glx and GABA following (es-)ketamine administration in A) healthy volunteers and B) MDD patients. Changes are depicted by symbols, with each symbol illustrating one finding. ↑ illustrates a significant increase in metabolite concentration; ^ illustrates a trend increase (0.05< p <0.10); ↓ illustrates a significant decrease in metabolite concentration; ៴ illustrates a trend decrease (0.05< p <0.10); = illustrates no changes in metabolite concentrations. dACC: dorsal anterior cingulate cortex; pgACC: pregenual anterior cingulate cortex; PCC: posterior cingulate cortex; OCC: occipital cortex.

#### 3.2.1 (Es-)Ketamine in healthy volunteers

Twelve studies recruited healthy volunteers to assess the effect of ketamine administration on metabolite concentrations. Of these studies, six investigated metabolite levels in the pgACC, five studies in the dACC, two studies in the PCC, and two studies in the hippocampus. These will be reported separately below (Table 2, Figure 3a). Studies varied with respect to the type of ketamine used, with 7 studies administering racemic ketamine (racemic mixture of both the R- and S-enantiomer of ketamine), 2 studies administering es-ketamine (only the S-enantiomer of ketamine), and 3 studies not specifying the type of ketamine used. Additionally, the infusion schemes were heterogeneous across studies, with some studies employing a single infusion and some separating the dosing scheme in a bolus and infusion. Doses reported in the text reflect the (estimated) total dose.

##### 3.2.1.1 Pregenual anterior cingulate cortex

The first study investigating the effect of intravenous racemic ketamine (0.5 mg/kg) on Glx, Glu and Glx/Glu was a placebo-controlled study, which did not report any significant effect between the ketamine (n=8) and placebo (n=9) groups, or a time effect of ketamine in the 40 minutes after infusion Taylor et al. 2012). Similarly, a study investigating Glx and GABA concentrations in 25 subjects found no significant changes in Glx, GABA or GABA/Glx 2h post-infusion of racemic ketamine (0.8 mg/kg) (Silberbauer et al., 2020). In contrast, Javitt et al. (2018) found an increase in Glx concentration in the pgACC in 31 subjects receiving ketamine compared to 16 subjects receiving placebo, but only in the first 15 minutes of the 60-minute racemic ketamine (0.66 mg/kg) infusion. Notably, the change from baseline in Glx over the entire 60-minute infusion period was not significantly different between treatment groups. Gärtner *et al*. (2022) also reported a trend increase in Glu, but no changes in Gln or Gln/Glu 25 minutes after es-ketamine infusion (∼0.4 mg/kg) in 17 subjects.

The only 7T study that assessed neurometabolism 1 hour post-infusion of racemic ketamine (0.5 mg/kg) found no change in Glu, Gln or Gln/Glu in the pgACC for both placebo (n=14) and ketamine (n=12) groups (Li et al., 2017). Interestingly, at the 24 hours post-infusion time point a significantly increased Gln/Glu ratio (but not Glu or Gln separately) was observed compared to baseline. This increase was also significantly larger than in the placebo group. In contrast, in a randomized, placebo-controlled crossover 7T study with 17 subjects, Evans *et al*. (2018) found no differences between racemic ketamine (0.5 mg/kg) and placebo infusions for Glu, Gln, and Gln/Glu 24h after ketamine administration. In contrast, another placebo-controlled 7T study observed higher Glu in the racemic ketamine (0.5 mg/kg; 40 subjects) group compared to the placebo group (40 subjects) 24 hours after administration (Colic et al., 2019).

##### 3.2.1.2 Dorsal anterior cingulate cortex

In addition to the pgACC voxel, Li et al. (2017) also assessed Glu, Gln and Gln/Glu at 1h post-infusion, and at 24 hours post-infusion in the dACC, but found no significant effects. Rowland et al. (2005) reported an increase in Gln, but not Glu, from baseline to a measurement during a bolus infusion of ketamine (up to ∼0.5 mg/kg; 10 participants); although this increase was not reported during maintenance infusion. In contrast, Stone et al. (2012) showed an increase in Glu, but not Gln or Glx after ketamine infusion (∼0.7 mg/kg in 13 subjects). Another study however, did not find any effects of es-ketamine (0.29 mg/kg) on dACC Glu, Gln and Glx, either after the bolus or in the maintenance phase in 25 subjects (Bojesen et al., 2018). Moreover, the only study using a MRSI reported no ketamine-induced alterations in Glx, GABA or GABA/Glx (Silberbauer et al., 2020).

##### 3.2.1.3 Posterior cingulate cortex

Bednarik et al. (2021) measured levels in Glu, Gln, Glx and Glu/Gln in the PCC of 12 participants before and 3h after a single racemic ketamine (0.5 mg/kg) infusion. No differences were observed between baseline and post-infusion measurements. The 3D-MRSI study of (Silberbauer et al. 2020) also found no effects of ketamine administration on Glx, GABA, or GABA/Glx.

##### 3.2.1.4 Hippocampus

Kraguljac et al. (2017) investigated the effect of racemic ketamine (∼0.5 mg/kg; 15 participants) on hippocampal Glx, which they measured during infusion. Compared to baseline, Glx levels during ketamine infusion were increased in the ketamine, but not in the placebo group. The 3D-MRSI study by Silberbauer et al. (2020) did not replicate this finding, but instead reported a reduction in hippocampal GABA.

##### 3.2.1.4 Other brain regions

GABA levels in the thalamus were not altered following ketamine infusion (Stone et al., 2012). Finally, Silberbauer et al. (2020) did not find effects of ketamine on the thalamus, insula or putamen.

#### 3.2.2 (Es-)Ketamine in MDD patients

In only three of the included studies, the effect of ketamine administration on metabolite levels in MDD patients was investigated. All studies included medication-free MDD patients (drug-free for at least 2 weeks) and administered a single infusion of 0.5 mg/kg racemic ketamine (Table 2 and Figure 3b).

##### 3.2.2.1 Pregenual anterior cingulate cortex

Milak et al. (2016) observed an increase in Glu during but not immediately after a 40-minute infusion in 11 MDD patients. They also investigated GABA levels, which were only increased 13 and 26 minutes after the start of the infusion. A 7T study by Evans et al. (2018) in 20 MDD patients observed a trend increase in Glu 24 hours after a ketamine infusion, but no changes in Gln or Glu/Gln.

##### 3.2.2.2 Occipital cortex

Valentine et al. (2011) assessed the effect of an *i.v.* infusion of racemic ketamine on Glu, Gln and GABA at 3 hours and 48 hours after the infusion. They did not observe an effect of ketamine on any of these metabolites at either time point.

#### 3.2.3 Relationship between (es-)ketamine administration or treatment and symptoms

Results from the study by Valentine et al. (2011) with 10 MDD patients indicated that changes in occipital Glu, GABA, and Gln content were not significantly correlated with changes in depressive symptoms after ketamine administration. This was observed for metabolite concentrations both at 3h and 48h after infusion. Additionally, baseline measures of glutamate, GABA and glutamine were not correlated with symptom severity scores at any time point. In the pgACC, Milak et al. (2016) reported that neither Glx nor GABA changes correlated with clinical response to ketamine in.

### 3.3 Risk of bias assessment of included studies

Of the included studies, 2 were regarded as both controlled intervention and before-after studies with no control group. Both quality assessment tools were completed for these studies.

The NIH Quality assessment tool *for controlled intervention studies* was conducted for 14 studies. Results from this assessment tool are visualized in Table S4 (Supplementary Results). Of the studies investigating the effect of SSRIs and SNRIs on neurometabolites, 2 studies were rated as high quality (i.e. low risk of bias), and 5 as fair quality (i.e. fair risk or bias). For the (es-)ketamine studies, 4 were rated as high quality, 2 as fair quality, and one study as poor quality (i.e. high risk of bias).

For a total of 27 studies, the NIH Quality assessment tool *for before-after studies with no control group* was used (Table S5, Supplementary Results). All 13 SSRI or SNRI studies were rated as high quality (i.e. low risk of bias), whereas for (es-)ketamine, 11 studies were rated as high quality (i.e. low risk of bias) and three as fair quality (i.e. fair risk of bias).

In addition, the MRS-Q tool was used to assess the quality of the ^1^H-MRS design. Outcomes of this tool are visualized for all included studies in Table S6 (Supplementary Results

## 4. DISCUSSION

In this systematic review, we investigated changes in glutamatergic and GABAergic metabolite levels, measured using ^1^H-MRS, in response to SSRIs, SNRIs, or (es-)ketamine. Studies investigating administration of, or treatment with SSRIs or SNRIs were generally underpowered, yielded widely varying results, with no consistent findings across voxel locations, specific populations or the investigated metabolite. For (es-)ketamine, the results are marginally more consistent, with results suggesting that (es-)ketamine increased glutamate levels in the pgACC and dACC at study-specific, but inconsistent, time points after administration only. However, as the majority of included studies reported no effect of either SSRIs, SNRIs, or (es-)ketamine on glutamatergic and GABAergic metabolite levels, the observed trend should be interpreted with caution.

### 4.1 Metabolite changes in response to SSRIs, SNRIs, and (es-)ketamine across brain regions

SSRIs, SNRIs, and (es-)ketamine have different neurotransmitter receptor targets that are differentially expressed throughout the brain, and may elicit therapeutic effects through different pathways. As such, it is conceivable that the brain region studied is an important contributor to the differential effects of these antidepressants on glutamatergic and GABAergic metabolites. Whereas some of the investigated brain regions may be directly affected by the AD medication, changes in other regions (e.g. the OCC) might be the result of medication-induced changes elsewhere. The pgACC is considered to be a critical brain region involved in the pathophysiology of MDD (Godlewska et al., 2018; Pizzagalli et al., 2018) and TRD (Salvadore et al., 2009; Salvadore and Zarate, 2010). As part of the limbic network, it plays a significant role in emotional processing (Pizzagalli, 2011). In MDD patients, the pgACC is thought to exhibit hyperactivity (Mayberg et al., 2005, 1999; Seminowicz et al., 2004), which is reduced after treatment with SSRIs (Drevets et al., 2002; Mayberg et al., 2000). Notably, both SSRIs (Arnone et al., 2018) and (es-)ketamine (Gärtner et al., 2022) increase connectivity between the pgACC and dorsolateral prefrontal cortex. Likewise, (es-)ketamine increases BOLD and cerebral blood flow, and alters connectivity in the dorsal part of the ACC (Bryant et al., 2019; De Simoni et al., 2013; Gärtner et al., 2022).

Therefore, the pgACC may be a key region in the shared biological pathway targeted by different types of antidepressant medication. Indeed, computational modeling suggests that glutamatergic disturbances in the pgACC (i.e. slower glutamate clearance) underlie systems-level alterations in the PFC in MDD (Ramirez-Mahaluf et al., 2017). They additionally showed that SSRIs, through serotonin (5-HT) 1A receptor-mediated hyperpolarization, could normalize these disturbances. (Es-)ketamine-related increases in PFC glutamate are supported by preclinical studies showing increased extracellular glutamate in rodents (Chowdhury et al., 2012) and results of a ^13^C-MRS study, which showed that (es-)ketamine increases glutamate-glutamine cycling in the medial PFC (Abdallah et al., 2018). Although this offers a potential shared neurobiological pathway through which SSRIs, SNRIs, and (es-)ketamine could exert their therapeutic effects, the results of our systematic search do not provide support for this (when measured with ^1^H-MRS).

For SSRIs and SNRIs, none of the studied brain regions yielded consistent results, suggesting that effects of these antidepressants either a) do not affect the glutamate or GABA system, b) effects on glutamate/GABA systems cannot readily be determined with ^1^H-MRS, c) effects are elicited in different (down-or upstream) brain regions, or d) the described studies lack statistical power to detect the effects of these antidepressants on metabolite levels. Moreover, the large heterogeneity observed in medication dose, route of administration and treatment duration, complicates interpretation of ^1^H-MRS findings following SSRI/SNRI administration. The (es-)ketamine studies showed less variation in the (es-)ketamine dose, treatment duration, and route of administration than studies investigating SSRIs or SNRIs. For these studies, a somewhat more consistent pattern emerges where findings show a tentative increase in glutamatergic neurometabolite levels in several subregions of the ACC, while fewer effects were observed in other brain regions such as the PCC and OCC.

Of note, from a methodological standpoint the quality of data obtained from different voxel locations may not be uniform. For instance, the OCC is not typically investigated in MDD but it can provide high-quality spectra with relative ease. Conversely, while the hippocampus and subgenual anterior cingulate cortex (sgACC) are widely regarded as important brain regions in the pathophysiology of MDD, they are notoriously difficult to shim and thus to acquire spectra with acceptable linewidths. In addition, the voxels from which ^1^H-MRS measurements are acquired are relatively large, thereby frequently including several smaller anatomical subregions. This is especially problematic in e.g. the anterior cingulate cortex, where different subregions have been shown to exhibit distinct expression patterns of glutamatergic receptor subtypes (Dou et al., 2013; Palomero-Gallagher et al., 2009). These methodological limitations must be taken into account when interpreting ^1^H-MRS studies.

### 4.2 Experimental design considerations across studies

For SSRIs and SNRIs, it is generally accepted that a treatment duration of several weeks is necessary to elicit therapeutic effects. We therefore investigated differences between acute, subchronic, or chronic SSRI/SNRI administrations. Differences in glutamatergic or GABAergic neurometabolism with varying treatment durations could be driven by adaptive processes at the 5-HT synapse in response to prolonged treatment (Sangkuhl et al., 2009), or through changes in brain-derived neurotrophic factor (BDNF)-driven neural plasticity (Björkholm and Monteggia, 2016). However, results of the included studies were generally conflicting; we therefore showed no evidence for an effect of treatment duration of SSRIs or SNRIs on glutamate/GABA levels. It should however be noted that most studies administering SSRIs and SNRIs were conducted in MDD patients, as part of treatment studies. Only one study investigated acute effects of SSRIs on glutamatergic or GABAergic metabolites, which impedes a full exploration of the effect of treatment duration in this review.

For (es-)ketamine, the effects of its administration on the brain over time are not as well understood. Although improvement in clinical symptoms occurs rapidly following (es-)ketamine infusion and is thought to sustain for several days, preclinical studies have shown both rapid and transient increases in glutamate release (a glutamate “burst”) following (es-)ketamine administration in the rat PFC (Moghaddam et al., 1997) and in humans (Homayoun and Moghaddam, 2007). Likewise, our findings show an apparently inconsistent effect of (es-)ketamine infusion over time on glutamate levels, with both significant effects and null findings observed shortly after the (es-)ketamine infusion, as well as 24 hours later. Many studies conducting multiple post-infusion measurements report changes at specific timepoints, which are frequently not sustained when averaging over the entire post-infusion period. However, as all (es-)ketamine studies used a single intravenous administration, predominantly in healthy volunteers, we were not able to determine long-term effects of (sub)chronic (es-)ketamine administration in the current work. Crucially, a full mechanistic explanation of the direct and indirect impact of (es-)ketamine on the human brain is lacking. Besides NMDA antagonism, (es-)ketamine has been shown to additionally affect multiple neurotransmitter systems, including the serotonergic (Gigliucci et al., 2013), noradrenergic (Tso et al., 2004), dopaminergic (Kokkinou et al., 2017), cholinergic (Moaddel et al., 2014) and opioid (Klein et al., 2020) systems. The interplay between these systems and (possibly subsequent) alterations in glutamate and GABA levels remains to be resolved.

### 4.3 Differences across diagnostic groups in metabolite changes

As previous studies have highlighted differences in glutamatergic and GABAergic metabolites in patients with MDD compared to healthy volunteers (Lener et al., 2017; Moriguchi et al., 2019; Yüksel and Öngür, 2010), we investigated the evidence for an effect of diagnostic group on the response to SSRIs, SNRIs and (es-)ketamine. For SSRIs and SNRIs, we observed no clear difference between the results in MDD patients *vs* controls. However, the consistency across all studies was minimal. Similarly, our results show that following (es-)ketamine administration both MDD patients and controls exhibit increases in glutamatergic metabolites in the anterior cingulate regions. However, these conclusions should be treated with caution due to lack of sufficient overlap of investigated brain regions between these two groups. We also cannot draw conclusions on how differences in in- and exclusion criteria affected the impact of antidepressants on neurometabolism, due to the small number of studies and variety in criteria. Nevertheless, it would be interesting for future studies to e.g. assess differences between patients with specific comorbidities, or those within specific subgroups of MDD, as es-ketamine is approved specifically for the treatment of TRD.

### 4.4 Association of metabolite levels with clinical response

Numerous ACC subregions are implicated in terms of predicting antidepressant treatment response. For example, studies have shown that response is predicted by the sgACC baseline metabolism (Nugent et al., 2014), pgACC gray matter volume (Chen et al., 2007), and pgACC reactivity (Strege et al., 2023) and is correlated with connectivity between the pgACC and dACC (Kozel et al., 2011).

For SSRIs and SNRIs, the findings show mixed results regarding the correlation between changes in Glu and GABA levels and clinical improvement in patients treated with SSRIs and SNRIs. Some, but not all studies found that increases in pgACC GABA levels were associated with clinical improvement. Notably, one study found that decreases in PCC Glu were associated with a reduction of depressive symptoms. However, in the OCC and dACC/dlPFC, no such association was evident. Our results furthermore suggest that changes in Glu, GABA, and Gln content are not significantly correlated with changes in depressive symptoms after (es-)ketamine administration, regardless of voxel placement. However, most included studies were conducted in small samples with considerable variation in clinical tools used to determine symptom severity. Additionally, it remains difficult to reliably determine the efficacy of treatment in these cohorts, particularly when employing treatment periods shorter than 6 weeks.

### 4.5 Study quality

Half of the studies on SSRIs and SNRIs used randomized, placebo-controlled designs, while the other half used an open-label design. Despite preference for a randomized placebo-controlled design, no differences were found between study designs. For (es-)ketamine studies, most used a placebo-controlled design, but conducting a blinded placebo-controlled study for (es-)ketamine can be challenging due to its potent subjective effects (Ingram et al., 2018). The quality of evidence from the included studies was generally fair to good, with many being of moderate risk of bias due to issues such as inadequate blinding or incomplete outcome data. Additionally, many of the included studies had small to moderate sample sizes and the majority did not show power calculations. Crucially, previous power analyses showed that a sample size of 18 subjects was necessary to detect a 10% difference in metabolite levels in a crossover design (Hansen et al., 2016). To detect a 15% change in GABA in the ACC a sample size of 27 participants would be necessary using a between-group design and 36 for a within-subject cross-over design (Sanaei Nezhad et al., 2019). Unfortunately, most studies had sample sizes well below these estimates and were therefore likely underpowered.

Considering ^1^H-MRS methodology, the studies in both healthy volunteers and MDD patients did not reveal any clear effect regarding the MRS sequence used. However, regarding field strength, we found moderate evidence that studies utilizing higher field strengths (7T) were more likely to detect an increase in Glu after (es-)ketamine administration in the pgACC. In contrast, studies that used lower field strengths (≤3T) seem to report no changes or only trends more frequently. Moreover, substantial differences in reporting of ^1^H-MRS acquisition, processing and analysis method existed between studies, which complicates an estimation of the quality and risk of bias of the included studies. Variation in reporting was most evident for the reference method used, the use of non open-access analysis tools and limitations in reporting of all software and processing steps used in data analysis. This highlights the importance to scan at higher field strengths, standardize acquisition protocols and increase transparency about analysis strategies to ensure the reliability and reproducibility of results across studies.

### 4.6 Future outlook

Several important areas to study the effects of antidepressants on the glutamate and GABA systems using ^1^H-MRS need to be considered. First, establishing standardized study and acquisition protocols and increasing transparency regarding analysis strategies is crucial to promote reliability and reproducibility. Additionally, measuring metabolites dynamically over time could provide valuable insights into the temporal dynamics of these systems (Lea-Carnall *et al.,* 2023; Mullins *et al*., 2005; Stanley & Raz, 2018), particularly as changes in metabolites in response to (es-)ketamine administration might occur at specific time points only (but not across the entire ^1^H-MRS acquisition period). A final, promising avenue is to combine MR spectroscopy with whole brain functional imaging techniques, such as functional MRI. This could provide a more complete picture of the relationship between metabolic activity and brain function (Bednařík et al., 2015; Ip et al., 2019; Koush et al., 2022).

### 4.7 Conclusion

Our systematic collection and review of ^1^H-MRS findings did not elucidate a clear effect of treatment with or administration of SSRI or SNRIs on GABAergic or glutamatergic metabolite levels in both healthy volunteers and MDD patients. Our findings suggest (es-)ketamine-induced alterations in glutamatergic metabolites might occur in the anterior cingulate regions in all subjects, but the timing of these effects remains to be resolved. Unfortunately, the many null findings due to generally underpowered studies complicate a reliable interpretation of these emerging trends. We did not find evidence for a comparable or shared effect of SSRIs and SNRIs, and (es-)ketamine on glutamatergic and GABAergic neurometabolism. The observed inconsistencies might (also) be related to differences in study design, the studied brain region, and ^1^H-MRS acquisition protocols and analysis approaches. Due to these differences, we were not able to conduct a meta-analysis of the combined evidence per brain region. Together, this highlights the need to standardize these aspects in future research. Additionally, as the response to these types of medication might be dynamic in nature, studies using functional ^1^H-MRS designs to assess both transient and sustained changes might elucidate the direct and downstream effects of these mechanistically distinct types of antidepressants. Finally, studies combining ^1^H-MRS with other imaging modalities, such as fMRI, could pave the way to understand the relation between changes in neurotransmission induced by these types of medication and its effect on the functional response of the brain.

## Declaration of Generative AI and AI-assisted technologies in the writing process

During the preparation of this work the author(s) used ChatGPT (OpenAI) in order to modify text. After using this tool, the author(s) reviewed and edited the content as needed and take(s) full responsibility for the content of the publication.

## Data Availability

Not applicable, the current study is a systematic review

