## Supplementary Materials for "Neurometabolite changes in response to antidepressant medication: a systematic review of ^1^H-MRS findings"

### *SUPPLEMENTARY METHODS*

**Table S1. Full search strategy and outcomes per search engine.**

| Search | | Search terms | Hits |
|---|---|---|---|
| **PubMed** | | | |
| | #1 | 1H-MRS[MeSH Terms] OR MRSI[tiab] OR MRS[tiab] OR ‘magnetic resonance spectroscopy’[MeSH Terms] OR "magnetic resonance spectroscop*"[tiab] OR "MR spectroscopy"[tiab] OR "MR spectroscopic"[tiab] OR metabolism[MeSH Terms] OR metabol*[tiab] OR neurometab*[tiab] OR neurochemistry[MeSH Terms] OR neurochem*[tiab] | 3,832,521 |
| | #2 | glutamate[MeSH Terms] OR glutamat*[tiab] OR GABA[MeSH Terms] OR GABA*[tiab] OR γ-aminobutyric*[tiab] OR gamma-aminobutyric*[tiab] OR glutamine*[tiab] | 264,245 |
| | #3 | ketamine[tiab] OR SNRI[MeSH Terms] OR SSRI[tiab] OR "selective serotonin reuptake inhibitor"[tiab] OR "selective serotonin reuptake inhibitors"[tiab] OR "selective serotonin and norepinephrine reuptake"[tiab] OR "selective serotonin-norepinephrine reuptake"[tiab] OR "selective serotonin-noradrenaline reuptake"[tiab] OR citalopram[tiab] OR escitalopram[tiab] OR fluoxetine[tiab] OR fluvoxamine[tiab] OR paroxetine[tiab] OR sertraline[tiab] OR dapoxetine[tiab] OR duloxetine[tiab] OR trazodone[tiab] OR venlafaxine[tiab] | 64,452 |
| | #4 | "Review"[Publication Type] OR "Systematic Review"[Publication Type] OR "Meta-Analysis"[Publication Type] OR "meta-analysis"[tiab] OR "systematic review"[tiab] OR "systematic literature review"[tiab] OR "Editorial"[Publication Type] OR "Comment"[Publication Type] | 4,711,206 |
| | #5 | animal[ti] OR rat[ti] OR rats[ti] OR mouse[ti] OR mice[ti] OR monkey*[ti] OR macaque*[ti] OR primate*[ti] OR rodent*[ti] OR dog[ti] OR dogs[ti] OR canine[ti] OR equine[ti] OR horse*[tiab] OR fish[ti] OR rabbit*[ti] | 2,038,208 |
| | **#1 AND #2 AND #3 NOT (#4 OR #5)** | | **334** |
| **Web of Science** | | | |
| | #1 | Ts= ((1H-MRS OR MRSI OR MRS OR "magnetic resonance spectroscopy" OR "magnetic resonance spectroscopic" OR “MR spectroscopy” OR “MR spectroscopic” OR metabol* OR neurometab* OR neurochem* )) | 1,848,662 |
| | #2 | Ts= (( glutamat* OR "GABA" OR GABA* OR γ-aminobutyric* OR gamma-aminobutyric* OR glutamine*)) | 294,209 |

| | | | |
|---|---|---|---|
| | #3 | TS=((*ketamine OR SNRI OR "selective serotonin and noradrenaline reuptake" OR "selective serotonin-noradrenaline reuptake" OR "selective serotonin and norepinephrine reuptake" OR "selective serotonin-norepinephrine reuptake" OR SSRI OR "selective serotonin reuptake" OR citalopram OR escitalopram OR fluoxetine OR fluvoxamine OR paroxetine OR sertraline OR dapoxetine OR duloxetine OR trazodone OR venlafaxine)) | 80,144 |
| | #4 | TS=(( "meta-analysis" OR "systematic review" OR "systematic literature review")) | 448,841 |
| | #5 | TI=((animal OR rat OR mouse] OR monkey OR macaque OR primate OR rodent OR dog OR canine OR equine OR horse OR fish OR rabbit)) | 2,472,223 |
| | **#1 AND #2 AND #3 NOT (#4 OR #5)** | | **416** |
| **Embase** | | | |
| | #1 | '1h-mrs'/exp OR '1h-mrs' OR mrsi:ti,ab,kw OR mrs:ti,ab,kw OR 'magnetic resonance spectroscopy'/exp OR 'magnetic resonance spectroscopy':ti,ab,kw OR 'magnetic resonance spectroscop*':ti,ab,kw OR 'mr spectroscop*':ti,ab,kw OR metabol*:ti,ab,kw OR neurometab*:ti,ab,kw OR neurochem*:ti,ab,kw | **2,133,473** |
| | #2 | 'glutamate'/exp OR 'glutamate' OR glutamate*:ti,ab,kw OR 'gaba'/exp OR gaba OR gaba*:ti,ab,kw OR 'γ aminobutyric*':ti,ab,kw OR glutamine*:ti,ab,kw | 355,600 |
| | #3 | 'ketamine'/exp OR ketamine:ti,ab,kw OR snri:ti,ab,kw OR ssri:ti,ab,kw OR 'selective serotonin reuptake inhibitor':ti,ab,kw OR 'selective serotonin reuptake inhibitor*':ti,ab,kw OR 'selective noradrenaline and serotonin reuptake':ti,ab,kw OR 'selective norepinephrine and serotonin reuptake':ti,ab,kw OR citalopram:ti,ab,kw OR escitalopram:ti,ab,kw OR fluoxetine:ti,ab,kw OR fluvoxamine:ti,ab,kw OR paroxetine:ti,ab,kw OR sertraline:ti,ab,kw OR dapoxetine:ti,ab,kw OR duloxetine:ti,ab,kw OR trazodone:ti,ab,kw OR venlafaxine:ti,ab,kw | 121,738 |
| | #5 | animal:ti OR rat:ti OR rats:ti OR mouse:ti OR mice:ti OR monkey*:ti OR macaque*:ti OR primate*:ti OR rodent*:ti OR dog:ti OR dogs:ti OR canine:ti OR equine:ti OR horse*:ti OR fish:ti OR rabbit*:ti | 2,372,993 |
| | **#1 AND #2 AND #3 NOT #4 AND ([article]/lim OR [article in press]/lim OR [letter]/lim)** | | **352** |

### *SUPPLEMENTARY RESULTS*

#### *S2 The effect of SSRIs and SNRIs on non-glutamatergic and non-GABAergic metabolites*

*S2.1 SSRIs and SNRIs in healthy volunteers*

Non-glutamatergic or -GABAergic metabolites that were investigated in the healthy volunteers were NAA (either tNAA (NAA+NAAG), NAA, or NAAG), (total) choline ((t)Cho), myo-inositol (mI), glutathione (GSH) and (total) creatine ((t)Cr). Two studies investigated the pgACC and did not observe an effect of SSRI or SNRI treatment on NAA or mI concentration (Hansen et al., 2016; Taylor et al., 2010) nor on Cho or Cr (Taylor et al., 2010). In the occipital cortex, Taylor et al. (2008) reported no treatment-induced changes in NAA, Cho, Cr, and mI. Maron et al. (2016) additionally reported a reduction in NAAG, but no changes in NAA or GSH. In the prefrontal cortex and the insula, no change in NAA or mI was observed following SNRI treatment (Hansen et al., 2016).

*S2.2 SSRIs and SNRIs in MDD patients*

Additionally investigated metabolites in MDD patients included (t)Cr, Cho, tNAA, NAA, NAAG, mI and GSH. In the pgACC, Taylor et al. (2012) reported an increase in NAA and tNAA in response to subchronic SSRI treatment, but no changes in Cho or mI. In a 1-year follow-up study, Draganov et al. (2020) investigated Cr, NAAG, mI, and GSH in the pgACC and observed an increase in mI, but not in any of the other metabolites. A trend decrease in dACC mI following citalopram treatment was observed in one study (Smith et al., 2021), but no changes in dACC NAA, NAAG, or GSH nor alterations in PCC NAA, NAA, GSH, or mI. No effect of citalopram treatment was observed on occipital GSH (Godlewska et al., 2015) nor on NAA, Cho, Cr or mI in the hippocampus (Block et al., 2009).

*S2.3 Relationship between metabolite measures and symptoms*

In the pgACC, one study conducting a one-year follow-up did not reveal significant relationships between changes in symptom severity and metabolite levels of NAAG, mI, Cr, or GSH over time (Draganov et al., 2020). In the hippocampus, Block et al. (2009) observed no correlation between baseline NAA, Cho, Cr, or mI with baseline symptom severity score. However, they reported a significant negative association between change in NAA and Cho and change in depressive symptoms, as well as a significant association between baseline NAA and Cho and change in symptom score. In the occipital cortex, Godlewska et al. (2015) found no correlation between change in the symptom severity and change in GSH level. Finally, Smith et al. (2021) found that increases in dACC GSH were associated with improvement in depressive symptoms.

#### *S3 The effect of (es-)ketamine on non-glutamatergic and -GABAergic metabolites*

*S3.1 (Es-)Ketamine in healthy volunteers*

Other metabolites that were investigated in the included articles were (t)NAA, (t)Cho, Asp, GSH, mI, Tau, Glc+tau, (t)Cr, and Glu/Asp. In the pgACC, no (es-)ketamine-induced differences in NAA, tCr, tCho, GSH or mI were reported (Bojesen et al., 2019; Evans et al., 2018). Likewise, in the dACC, no changes in NAA, Cho and Cr were observed by Rowland et al. (2005). In the PCC, no changes were reported in tNAA, Cho, Asp, GSH, mI, Tau, Glc+tau, tCr and Glu/Asp (Bednarik et al. 2017). Lastly, Kraguljac et al. (2017) revealed no racemic ketamine-induced alterations in hippocampal tNAA and tCho.

*S3.2 (Es-)Ketamine in MDD patients*

Evans et al. (2018) observed a trend increase in tNAA in the pgACC from baseline to post-infusion for both racemic ketamine and placebo, but reported no racemic ketamine-induced changes in GSH or tCho. In the OCC, Valentine et al. (2011) also reported no changes in tNAA, tCho, tCr, and mI at 3h or 48h after a racemic ketamine infusion.

Table S2. Outcomes of included studies investigating the effect of SSRI or SNRI administration on non-glutamatergic and non-GABAergic metabolites.

| Article | Sample | Design | Medication | Dose | Duration | $^{1}$H-MRS timepoint | Field strength and $^{1}$H-MRS sequence | VOI and voxel size (mm) | Metabolites | Quantification method | Results |
|---|---|---|---|---|---|---|---|---|---|---|---|
| **HC** | | | | | | | | | | | |
| Hansen et al. (2016) | HC<br>N=20 | randomized, placebo-controlled, double-blind crossover design | Oral venlafaxine | 75 mg/day | 5 days | Post-treatment | 3T PRESS | pgACC<br>20 x 20 x 20<br><br>Insula<br>15 x 20 x 50<br><br>PFC<br>15 x 25 x 20 | NAA, mI | LCModel | = NAA and mI in pgACC, PFC and insula between venlafaxine and placebo |
| Maron et al. (2016) | HC<br>N=15 | open-label pre-post design without placebo | Oral escitalopram | 10 mg/day | 7-10 days | Post-treatment | 3T SPECIAL | OCC<br>20 x 25 x 20 | GSH, NAA, NAAG | LCModel | = GSH and NAA between baseline and post-treatment<br>↓ NAAG from baseline to post-treatment |
| Taylor et al. (2008) | HC<br>N=30 | randomized, placebo-controlled, parallel group design | Oral citalopram<br><br>Oral Reboxetine | 20 mg/day<br><br>8 mg/day | 7-10 days | Post-treatment | 3T PRESS | OCC<br>n.s. | NAA, Cho, Cr, mI | LCModel | = NAA, Cho, Cr and mI post-treatment between treatment groups |
| Taylor et al. (2010) | HC<br>N=23 SSRI<br>N=10 PLAC | randomized, placebo-controlled, parallel group design | Oral citalopram | 20 mg/day | 7-10 days | Post-treatment | 3T PRESS<br><br>3T J-PRESS | pgACC<br>20 x 20 x 20 | NAA, Cho, Cr, mI | LCModel | = NAA, Cho, Cr and mI between baseline and post-treatment across groups |

| | | | | | | | | | | | |
|---|---|---|---|---|---|---|---|---|---|---|---|
| **MDD** | | | | | | | | | | | |
| Block et al. (2009) | MDD<br>N=5 SSRI<br><br>N=5 TCA | pre-post design without placebo | Oral citalopram<br><br>Oral nortriptyline | Mean 20 mg/day<br><br>Mean 105 mg/day | 8 weeks | Post-treatment | 3T PRESS | HIPP<br>28 x 17 x 13 | NAA, Cho, Cr, mI | AMARES | = NAA, Cho, Cr, and mI between baseline and post-treatment<br>No differences between groups<br>No correlation between baseline NAA, Cho, Cr, or mI and baseline BDI<br>Correlation between ΔCho and ΔNAA and ΔBDI<br>No association between ΔCr or ΔIns and ΔBDI |
| Draganov et al. (2020) | MDD<br>N=18 | open-label pre-post design without placebo | Variable, mostly oral escitalopram | Mostly 15 mg/day | 1 year | Post-treatment | 3T PRESS | pgACC<br>20 x 20 x 20 | NAAG, mI, Cr, GSH | TARQUIN | ↑ mI from baseline to post-treatment<br>= NAAG, Cr and GSH between baseline and post-treatment<br>No correlation between ΔCr, ΔNAAG, ΔIns or ΔGSH and ΔHDRS |
| Godlewska et al. (2015) | MDD<br>N=39 | open-label pre-post design without placebo | Oral citalopram | 10 mg/day | 6 weeks | Post-treatment | 3T SPECIAL | OCC<br>20 x 25 x 20 | GSH | LCModel | = GSH between baseline and post-treatment<br>No correlation between ΔHAM-D and ΔGSH |
| Taylor et al. (2012) | MDD<br>N=21 SSRI<br>N=19 PLAC | randomized, double-blind parallel group design | Oral escitalopram | 10 mg/day | 7 days | Day 7 | 3T PRESS | pgACC<br>30 x 3- x 20 | NAA, tNAA, Cho, mI | LCModel | ↑ NAA and tNAA in treatment group compared to placebo group<br>= Cho and mI between treatment group and placebo group |

| | | | | | | | | | | | |
|---|---|---|---|---|---|---|---|---|---|---|---|
| Smith et al. (2021) | MDD<br>N=9 | open-label pre-post design without placebo | Oral citalopram | w1<br>10 mg/day<br>w2-12<br>20-40 mg/day | 10-12 weeks | Post-treatment | 7T STEAM | dACC<br>28 x 20 x 16<br><br>PCC<br>28 x 20 x 16 | NAA, NAAG, GSH, mI | LCModel | ∨ dACC mI from baseline to post-treatment<br>= NAA and GSH in dACC between baseline and post-treatment<br>= NAA, GSH, and mI in PCC between baseline and post-treatment<br>Association between dACC ΔGSH and ΔBDI |

Abbreviations: BDI: Beck Depression Inventory; (t)Cho: (total) choline; (t)Cr: (total) creatine; dACC: dorsal anterior cingulate cortex; Glu: glutamate; Gln: glutamine; Glx: glutamate+glutamine; GABA: γ-aminobutyric acid; GSH: glutathione; HC: healthy volunteers; HDRS: Hamilton Depression Rating Scale; MADRS: Montgomery-Asberg Depression Rating Scale; MDD: Major Depressive Disorder; mI: myo-inositol; (t)NAA: (total) N-acetylaspartate; NAAG: N-acetyl-aspartyl-glutamate; n.s.: not specified; OCC: occipital cortex; PCC: posterior cingulate cortex; pgACC: pregenual anterior cingulate cortex; PFC: prefrontal cortex; PLAC: placebo; SNRI: serotonin and noradrenaline reuptake inhibitor; SSRI: selective serotonin reuptake inhibitor; TCA: tricyclic antidepressant; tCr: total creatine.

**Table S3. Outcomes of included studies investigating the effect of (es-)ketamine administration on non-glutamatergic and non-GABAergic metabolites**

| Article | Sample | Design | Medication | Dose | Duration | $^{1}$H-MRS timepoint | Field strength and $^{1}$H-MRS sequence | VOI and voxel size (mm) | Metabolites | Quantification method | Results |
|---|---|---|---|---|---|---|---|---|---|---|---|
| **HC** | | | | | | | | | | | |
| Bednarik et al. (2021) | HC N=12 | open-label pre-post design without placebo | IV Racemic ketamine | 0.8 mg/kg | 50 min. | 3h post-infusion | 3T sLASER | PCC 22 x 22 x 22 | Asp, GSH, mI, Tau, Glx+Tau, tCho, tCre, Glu/Asp | LCModel | = Asp, GSH, mI, Tau, Glx+Tau, tCho, tCre, Glu/Asp from baseline to post-infusion |
| Bojesen et al. (2019) | HC N=25 | open-label pre-post design without placebo | IV Es-ketamine | 0.25 mg/kg + 0.125 mg/kg | 20 min + 20 min | During bolus 2x during infusion | 3T PRESS | pgACC 20 x 20 x 20 | NAA, tCr, tCho, mI | LCModel | = NAA, tCr, tCho, mI between baseline and during bolus or during infusion |
| Evans et al. (2018) | HC N=17 | randomized, placebo-controlled, double-blind crossover design | IV Racemic ketamine | 0.5 mg/kg | n.s. | 24h post-infusion | 7T PRESS | pgACC 20 x 20 x 20 | GSH, tNAA, Cho | In-house linear combination | = GSH, tNAA, tCho between baseline and post-administration |
| Kraguljac et al. (2017) | HC N=15 | placebo-controlled crossover design | IV Racemic ketamine | 0.27 mg/kg + 0.25 mg/kg/h | 10 min + d.s. | 13 min. post start infusion | 3T PRESS | HIPP 26 x 15 x 10 | tNAA, tCho | AMARES | = tNAA and tCho between baseline and during infusion |

| | | | | | | | | | | | |
|---|---|---|---|---|---|---|---|---|---|---|---|
| Rowland et al. (2005) | HC N=10 | placebo-controlled crossover design | IV n.s. | 0.27 mg/kg + 0.135 mg/kg/h | 20 min + d.s. | During bolus Start infusion | 4T STEAM | dACC 8 mL | NAA, Cho, Cr | n.s. | = NAA, Cho, and Cr between baseline and during bolus or at start of infusion |
| **MDD** | | | | | | | | | | | |
| Evans et al. (2018) | HC N=17 | | IV Racemic ketamine | 0.5 mg/kg | n.s. | 24h post-infusion | 7T J-PRESS | pgACC 20 x 20 x 20 | GSH, tNAA, tCho | In-house linear combination | ^ tNAA from baseline to post-infusion in both ketamine and placebo groups<br>= GSH and tCho between baseline and post-administration |
| Valentine et al. (2011) | MDD N=10 | | IV Racemic ketamine | 0.5 mg/kg | 40 min. | 3h post-infusion 48h post-infusion | 4T J-edited sequence | OCC 30 x 15 x 30 | tNAA, tCho, tCr, mI | LCModel | = tNAA, tCho, tCr, and mI from baseline to all 3h or 48h post-infusion |

Abbreviations: Asp: aspartate; BDI: Beck Depression Inventory; (t)Cho: (total) choline; (t)Cr: (total) creatine; dACC: dorsal anterior cingulate cortex; d.s.: duration of scan; GABA: γ-aminobutyric acid; Glu: glutamate; Gln: glutamine; Glx: glutamate+glutamine; GSH: glutathione; i.v.: intravenous; pgACC: pregenual anterior cingulate cortex; HC: healthy volunteers; HDRS: Hamilton Depression Rating Scale; KET: ketamine; MADRS: Montgomery-Asberg Depression Rating Scale; MDD: Major Depressive Disorder; mI: myo-inositol; (t)NAA: (total) N-acetylaspartate; NAAG: N-acetyl-aspartyl-glutamate; n.s.: not specified; OCC: occipital cortex; PCC: posterior cingulate cortex; PFC: prefrontal cortex; PLAC: placebo; POMS: Profile of Mood States.

**Table S2. Outcomes of NIH quality assessment for controlled intervention studies.** Results are depicted as individual components per question of the quality assessment tool as well as the overall quality rating. In the used color scheme, green indicates a high quality (i.e. low risk of bias), yellow indicating a fair quality (i.e. fair risk of bias), and red indicating a poor quality (i.e. high risk of bias). Grey squares are depicted when a question was rated as not applicable.

| Article | Q1 | Q2 | Q3 | Q4 | Q5 | Q6 | Q7 | Q8 | Q9 | Q10 | Q11 | Q12 | Q13 | Q14 | Overall |
|---|---|---|---|---|---|---|---|---|---|---|---|---|---|---|---|
| **SSRI/SNRI** | | | | | | | | | | | | | | | |
| Bhagwagar et al. (2004) | | CD | | | NR | | | | | | | CD | CD | | |
| Hansen et al. (2016) | | CD | | | NR | | | | NR | NR | | | CD | | |
| Narayan et al. (2022) | | | | | | | | NR | NR | NR | | CD | CD | | |
| Spurny et al. (2021) | | CD | | | NR | NR | NR | NR | NR | | | CD | CD | | |
| Taylor et al. (2008) | | CD | | | NR | | | | NR | NR | | CD | CD | | |
| Taylor et al. (2010) | | CD | NR | NR | NR | | | | NR | NR | | CD | CD | | |
| Taylor et al. (2012) | | CD | | | NR | | NR | NR | NR | NR | | CD | CD | | |
| **(Es-)ketamine** | | | | | | | | | | | | | | | |
| Colic et al. (2019) | | | | | NR | | | | | | | CD | CD | | |
| Evans et al. (2018) | | CD | | | NR | | | | | | | CD | | | |
| Javitt et al. (2017) | | | | NR | NR | NR | | | | | | CD | CD | | |
| Li et al. (2017) | | | | | NR | | | | | | | | CD | | |
| Rowland et al. (2005) | | | | | NR | | | | | NR | | CD | CD | | |
| Taylor et al. (2012) | | CD | NR | NR | | | | | | | | CD | CD | | |
| Valentine et al. (2011) | | | | NR | | | | | | | | CD | CD | | |

Abbreviations: CD: cannot determine; NR: not reported; Q: question.

**Table S3. Outcomes of NIH quality assessment for before-after studies with no control group.** Results are depicted as individual components per question of the quality assessment tool as well as the overall quality rating. In the used color scheme, green indicates a high quality (i.e. low risk of bias), yellow indicating a fair quality (i.e. fair risk of bias), and red indicating a poor quality (i.e. high risk of bias). Grey squares are depicted when a question was rated as not applicable.

| **Article** | **Q1** | **Q2** | **Q3** | **Q4** | **Q5** | **Q6** | **Q7** | **Q8** | **Q9** | **Q10** | **Q11** | **Q12** | **Overall** |
|---|---|---|---|---|---|---|---|---|---|---|---|---|---|
| **SSRI/SNRI** | | | | | | | | | | | | | |
| Bhagwagar et al. (2004) | | | | | CD | | | NR | | | | | |
| Block et al. (2009) | | | | | CD | | | NR | | | | | |
| Brennan et al. (2017) | | | | | CD | | | NR | | | | | |
| Draganov et al. (2020) | | | | | CD | | | | | | | | |
| Godlweska et al. (2015) | | | | | CD | | | NR | NR | | | | |
| Grimm et al. (2012) | | | | | CD | | | NR | NR | | | | |
| Hansen et al. (2016) | | | | | | | | NR | | | | | |
| Maron et al. (2016) | | | | | CD | | | NR | NR | | | | |
| Narayan et al. (2022) | | | | | CD | | | | | | | | |
| Sanacora et al. (2002) | | | | | CD | | | NR | | | | | |
| Smith et al. (2021) | | | | | | | | | | | | | |
| Spurny et al. (2021) | | | | | CD | | | NR | NR | | | | |
| Taylor et al. (2010) | | | | | CD | | | NR | | | | | |
| **(Es-)ketamine** | | | | | | | | | | | | | |
| Bednarik et al. (2021) | | | | NR | CD | | | NR | NR | | | | |
| Bojesen et al. (2019) | | | | | CD | | | NR | | | | | |
| Colic et al. (2019) | | | | | CD | | | NR | | | | | |
| Evans et al. (2018) | | | | | CD | | | NR | | | | | |
| Gartner et al. (2022) | | | | | CD | | | NR | CD | | | | |
| Javitt et al. (2017) | | | | | CD | | | NR | | | | | |
| Kraguljac et al. (2017) | | | | | CD | | | | | | | | |
| Li et al. (2017) | | | | | CD | | | NR | | | | | |
| Milak et al. (2016) | | | | | CD | | | NR | | | | | |
| Rowland et al. (2005) | | | | | CD | | | NR | | | | | |
| Silberbauer et al. (2021) | | | | | CD | | | NR | | | | | |
| Stone et al. (2012) | | | | | CD | | | NR | | | | | |
| Taylor et al. (2012) | | | | | CD | | | NR | | | | | |
| Valentine et al. (2011) | | | | | CD | | | | | | | | |

Abbreviations: CD: cannot determine; NR: not reported; Q: question.

**Table S4. Outcomes of the MRS-Q assessment tool.** Results are depicted as individual components per question of the quality assessment tool. In the used color scheme, green indicates that the component above is satisfied, yellow a component is partially satisfied, and red indicates a component is not satisfied. Grey squares are depicted when the component was rated as not applicable.

| | | Parameters | | | Quality metrics | | | Study design/analysis | | | |
|---|---|---|---|---|---|---|---|---|---|---|---|
| **Reference** | **Scanner strength** | **Sequence** | **Parameters** | **Data points** | **Quality measures** | **Data visualization** | **Scanner drift** | **Power calculation** | **Partial volume correction** | **Frequency and phase correction** | **Analysis** |
| **SSRI/SNRI** | | | | | | | | | | | |
| **HC** | | | | | | | | | | | |
| Bhagwagar et al. (2004) | | | | | | | | | | | |
| Hansen et al. (2016) | | | | | | | | | | | |
| Maron et al. (2016) | | | | | | | | | | | |
| Spurny et al. (2021) | | | | | | | | | | | |
| Taylor et al. (2008) | | | | | | | | | | | |
| | | GABA | GABA | GABA | GABA | | | | | | |
| Taylor et al. (2010) | | | | | | | | | | | |
| **MDD** | | | | | | | | | | | |
| Block et al. (2009) | | | | | | | | | | | |
| Brennan et al. (2017) | | | | | | | | | | | |
| | | GABA | GABA | GABA | | | | | | | |
| Draganov et al. (2020) | | | | | | | | | | | |
| | | GABA | GABA | GABA | | | | | | | |
| Godlweska et al. (2015) | | | | | | | | | | | |
| Grimm et al. (2012) | | | | | | | | | | | |
| Narayan et al. (2022) | | | | | | | | | | | |
| Sanacora et al. (2002) | | | | | | | | | | | |
| Smith et al. (2021) | | | | | | | | | | | |
| Taylor et al. (2012) | | | | | | | | | | | |
| **Ketamine** | | | | | | | | | | | |
| **HC** | | | | | | | | | | | |
| Bednarik et al. (2021) | | | | | | | | | | | |
| Bojesen et al. (2019) | | | | | | | | | | | |
| Colic et al. (2019) | | | | | | | | | | | |

| | | | | | | | | | | | |
|---|---|---|---|---|---|---|---|---|---|---|---|
| Evans et al. (2018) | | | | | | | | | | | |
| Gartner et al. (2022) | | | | | | | | | | | |
| Javitt et al. (2017) | | | | | | | | | | | |
| Kraguljac et al. (2017) | | | | | | | | | | | |
| Li et al. (2017) | | | | | | | | | | | |
| Rowland et al. (2005) | | | | | | | | | | | |
| Silberbauer et al. (2021) | | | | | | | | | | | |
| Stone et al. (2012) | | | | GABA | GABA | | | | | | GABA |
| Taylor et al. (2012) | | | | | | | | | | | |
| **MDD** | | | | | | | | | | | |
| Evans et al. (2018) | | | | | | | | | | | |
| Milak et al. (2016) | | | | | | | | | | | |
| Valentine et al. (2011) | | | | | | | | | | | |
